# Network Diffusion Embedding Reveals Transdiagnostic Subnetwork Disruption and Potential Treatment Targets in Internalizing Psychopathologies

**DOI:** 10.1101/2021.04.01.21254790

**Authors:** Paul J. Thomas, Alex Leow, Heide Klumpp, K. Luan Phan, Olusola Ajilore

## Abstract

Network diffusion models are a common and powerful way to study the propagation of information through a complex system and they offer straightforward approaches for studying multimodal brain network data. We developed an analytic framework to identify brain subnetworks with perturbed information diffusion capacity using the structural basis that best maps to resting state functional connectivity and applied it towards a heterogenous dataset of internalizing psychopathologies (IPs), a set of psychiatric conditions in which similar brain network deficits are found across the swath of the disorders, but a unifying neuropathological substrate for transdiagnostic symptom expression is currently unknown. This research provides preliminary evidence of a transdiagnostic brain subnetwork deficit characterized by information diffusion impairment of the right area 8BM, a key brain region involved in organizing a broad spectrum of cognitive tasks, that may underlie previously reported dysfunction of multiple brain circuits in the IPs. We also demonstrate that models of neuromodulation involving targeting this brain region normalize IP diffusion dynamics towards those of healthy controls. These analyses provide a framework for multimodal methods that identify both brain subnetworks with disrupted information diffusion and potential targets of these subnetworks for therapeutic neuromodulatory intervention based on previously well-characterized methodology.

## INTRODUCTION

The dynamics arising from the interaction of individual elements of a complex system are commonly investigated by representing such a system as a network or graph (Bullmore and Sporns, 2009). This has been applied extensively towards studying neural connectivity, where brain regions (represented by nodes) and the links between them (represented by edges) define a brain network, or connectome (Sporns, 2010, 2018). As in any network, the underlying structure of connectivity constrains the functions and processes that take place within it (Wang et al., 2017). Indeed, the mechanisms of propagation of and neural response to brain pathologies have been closely tied to network topology (Fornito et al., 2015).

A number of studies have found that the basis formed by the *anatomic* structure of the brain and the patterns of observed spatiotemporal neural activity are intimately related (Atasoy et al., 2016; Deslauriers-Gauthier et al., 2020; Abdelnour et al., 2014, 2018). Many of these investigations use a network diffusion model to study the propagation of neural impulses, or information (given by functional connectivity), throughout the structure formed by the white matter tracts of the brain. These so-called network diffusion-based approaches all utilize the properties of the structural graph Laplacian, which encodes how a diffusive process spreads throughout a network over time (Chung and Graham, 1997). The basis of diffusion given by the graph Laplacian, or its eigenmodes, can thus be used to model the flow of neural information (Xiao et al., 2010), and has been shown to play important roles in healthy brain network organization (Wang et al., 2017). More specifically, such an approach was used successfully to demonstrate that resting state functional connectivity is predictable from white matter structural eigenmodes (Abdelnour et al., 2014, 2018). Furthermore, resting state networks were found to closely match spatial patterns of structural eigenmodes (Atasoy et al., 2016). Of note, many other approaches for investigating this interplay have been studied, including models based on epidemic-spreading (Stam et al., 2016), threshold models (Mišić et al., 2015) and neural mass models (Honey et al., 2009) (for a review, see (Avena-Koenigsberger et al., 2018)).

Network diffusion-based methods have also been applied towards investigating the pathophysiology of neurological diseases. For example, Raj and colleagues (Raj et al., 2012) show that white matter structural eigenmodes closely resemble known patterns of dementia and correlate strongly with regional atrophy. A closely related methodology to that of studying structural eigenmodes is using the graph Laplacian-derived heat kernel of brain networks, a matrix which encodes how much information is transferred between every pair of network nodes after a given time, or diffusion depth. Heat kernel methods have been used to characterize perturbations in brain network information transfer in autism (Schirmer and Chung, 2019) and to predict future adverse motor function resulting from premature birth using white matter structural connectomes (Chung et al., 2016).

In the context of psychiatric conditions, the application of methods that integrate multimodal perspectives in their analytical approaches has great potential to elucidate the distributed perturbation of neurocircuitry that underlie the complex cognitive and behavioral disruption found in patients suffering from these disorders. This may hold particularly true for the internalizing psychopathologies (IPs), including mood (e.g., major depressive disorder (MDD), dysthymia) and anxiety (e.g., panic disorder (PD), social anxiety disorder (SAD), generalized anxiety disorder (GAD)) disorders. Many previous neuroimaging studies have been conducted on IPs, however, most of which use unimodal data analysis. Importantly, findings about potential neuropathological substrates are more often overlapping between the IPs than distinct to a specific IP. In addition, IPs, as traditionally categorized, are often comorbid with one another and present heterogeneously with a spectrum of related symptoms and disruptions to emotion regulation and negative valence system (NVS) processes (Kessler et al., 2005; Moser et al., 2015; Cecilione et al., 2018). Furthermore, the available first line treatments for the IPs, either cognitive behavioral therapy (CBT) or selective serotonin reuptake inhibitors (SSRI), are equally effective across the swath of the disorders (Dunlop et al., 2012). Many studies have concluded that similar structural and functional brain networks involving regions commonly implicated in the expression of fear, anxiety, negative affect and other NVS features are dysfunctional in these disorders (Etkin and Wager, 2007; Hamilton et al., 2012; Korgaonkar et al., 2014). In addition, previous findings also implicate the disruption of similar canonical resting state networks (RSNs), such as the default mode network (DMN), in both depression and anxiety using both structural and functional neuroimaging (Müller et al., 2013; Sheline et al., 2009; Liu et al., 2020; Tao et al., 2015; Kim and Yoon, 2018). To address this pattern of findings, the National Institute of Mental Health’s Research Domain Criteria (RDoC) initiative was developed (Insel et al., 2010; Cuthbert, 2014) in order to reorient the study of psychiatric disorders away from traditional diagnostic categories and towards that of data-driven approaches such as identifying transdiagnostic cognitive domain disruptions, biomarkers of treatment response and targets for neuromodulatory intervention.

In line with the aims set forth by this initiative, we study a dataset from an RDoC clinical trial consisting of diffusion weighted (DWI) and resting state-functional (rs-fMRI) neuroimaging scans of a treatment naive, heterogeneous IP patient (PT) cohort and age and sex matched healthy controls (HC). PTs were then randomized to either 12 weeks of SSRI or CBT and completed Inventory of Depression and Anxiety Symptoms (IDAS-II) (Watson et al., 2012) self-reports pre- (Pre) and post-treatment (Post) to assess transdiagnostic dimensions of symptom burden. Several previous analyses have been conducted on this dataset but they are unimodal and have a hypothesis-driven focus on *a priori* brain region, often studied in association with specific task-based measures of NVS subsystems (Radoman et al., 2019; Burkhouse et al., 2018a, 2020; Gorka et al., 2019; Thomas et al., 2020; Klumpp et al., 2020).

Because IPs share similarly dysfunctional brain networks and respond to similar treatments, we hypothesize that there exist pathophysiological features common to all IPs that result in impaired emotion regulation and the subsequent heterogeneous expression of IP symptoms. In this paper we ask (1) can the presence of such unifying perturbations can be identified by incorporating both structural and functional connectivity data, (2) do these perturbations predict response to CBT and/or SSRI treatment and (3) can we identify candidate brain region as targets of neuromodulatory intervention to normalize connectivity dynamics to those found in healthy controls? To answer these questions, we use a multimodal data-driven analysis based on network diffusion models. In this approach, the diffusion basis of the structural connectome, given by the eigenmodes (eigenvector-eigenvalue pairs) of the normalized graph Laplacian, is central to the methodology. The eigenmode basis of a network encodes its information diffusion properties, and the eigenmodes of a graph, scaled exponentially by the diffusion depth or time scale allowed for diffusion to occur, represent an embedding of nodes such that their pairwise Euclidean distance in the embedding is inversely proportional to the ability for information to diffuse between nodes at the given diffusion depth (Chung et al., 2016; Xiao et al., 2010, 2005). Next, the mapping between each subject’s structural and functional network that minimizes the error between the empirical and estimated functional connectivity is computed by optimizing over diffusion depth parameters as described by Abdelnour and colleagues (Abdelnour et al., 2018). We then define a novel representation of structural connectivity, the structural diffusion distance (SDD) connectome, where the edge weights between each brain network node are given by the pairwise Euclidean distance from the diffusion embedding at the optimal time scale that best maps to empirical rs-fMRI activity. We chose such a model for multimodal incorporation as it has both been well characterized in the study of structural to functional mappings in brain networks (Abdelnour et al., 2018) and allows for the computation of the diffusion-based network embedding given by the structural eigenbasis that most accurately underlies empirical functional connectivity. Thus, we achieve correspondence to functional connectivity in SDD connectomes by computing the embedding at the optimal diffusion depth, *βt*, for each subject (see Methods section for details). Furthermore, a network diffusion model easily allows for the simulation of information flowing through the network. Simple mathematical properties of the graph Laplacian-derived heat kernel corresponding to the network embedding can be exploited to identify network nodes to supplement with additional ‘heat’ supplement in order to normalize deficient heat transfer throughout a PT subnetwork towards the desired diffusion dynamics of a target (mean HC) subnetwork. Our study applies this analytic framework to provide evidence for transdiagnostic IP subnetwork disruption and neuromodulation treatment targets, opening up application of multimodal network diffusion-based methods towards other brain disorders.

## MATERIALS AND METHODS

### Clinical trial and research participants

Subjects were recruited from the greater Chicago area through advertisements and through University of Illinois at Chicago (UIC) outpatient clinics and counselling centers as part of a larger Research Domain Criteria (RDoC) (Cuthbert, 2014) investigation on predictors of IP treatment outcomes (ClinicalTrials.gov identifier: NCT01903447). A heterogeneous study population was recruited in order to obtain a sample with a broad range of symptom severity and functioning. Details regarding inclusion/exclusion criteria, participant recruitment, clinical characteristics and treatment have been previously described (Gorka et al., 2019). In brief, this study was approved by the UIC Institutional Review Board, and written informed consent was obtained for each participant. The inclusion criteria for subjects were age between 18 and 65 years, and the need for randomization to 12 weeks of treatment with SSRI or CBT, as determined by a consensus panel consisting of at least three trained clinicians or study staff. Subjects were excluded from the study if they have a history of current or past manic/hypomanic episodes or psychotic symptoms, active suicidal ideation, presence of contraindications or history of SSRI resistance (no response to >2 SSRIs despite adequate duration and dose), psychopathology not appropriate for the treatment algorithm, or current cognitive dysfunction or impairment. The SCID-5 (First et al., 2015) was used to determine current and lifetime Axis I diagnoses. The study was a parallel group randomized control trial with 1:1 allocation ratio to either 12 weeks of CBT or SSRI. For the SSRI cohort, PTs were administered one of 5 drugs (sertraline, fluoxetine, paroxetine, escitalopram or citalopram) with a flexible dosing schedule with a goal of obtaining target dose by 8 weeks. SSRI PTs met at 0, 2, 4, 8 and 12 weeks with their study psychiatrist for medication management. For the CBT cohort, PTs received 12 once-weekly 60 min sessions led by a PhD-level clinical psychologist. CBT procedures were based on the PT’s principal diagnosis and predominant symptoms (Burkhouse et al., 2020). Each participant was scanned at enrollment and IP subject scans were acquired before treatment was administered.

At the time of enrollment (Pre) and after 12 weeks of treatment (Post), severity of IP symptoms was assessed in all subjects using the Inventory of Depression and Anxiety Symptoms (IDAS-II) (Watson et al., 2012). Subjects responded to each of the 99 items in this inventory using a 5-point Likert-type scale ranging from 1 (not at all) to 5 (extremely), yielding 17 empirically derived and symptom-specific scales (Suicidality, Lassitude, Insomnia, Appetite Loss, Appetite Gain, Ill-Temper, Well-Being, Panic, Social Anxiety, Traumatic Intrusions, Traumatic Avoidance, Mania, Euphoria, Claustrophobia, Checking, Ordering, Cleaning, General Depression and Dysphoria). As we are interested in transdiagnostic NVS construct disruptions in IPs, we use the IDAS-II Panic and Depression subscales, as these scales have been shown to map well to ‘fear’ and ‘distress’ dimensions, respectively, which is a previously used approach for broadly dividing and assessing these symptom domains (Radoman et al., 2019; Watson et al., 2012; Ofrat and Krueger, 2012).

### Image acquisition and processing

All imaging was acquired at the UIC Center for Magnetic Resonance Research using a 3 Tesla GE Discovery MR750 System (Milwaukee, WI) with an 8-channel head coil.

#### Anatomic MRI

High resolution 1 mm isotropic voxel resolution T1-weighted (T1w) images were obtained using a 3D axial FSPGR BRAVO imaging sequence with the following parameters: slice thickness = 1 mm, in-plane resolution = 1 mm, repetition time (TR) = 9.3 ms, echo time (TE) = 3.8 ms, inversion time (TI) = 450 ms, flip angle = 13°, field of view (FOV) = 220 × 220 mm.

#### Diffusion weighted MRI

Diffusion weighted images (DWI) were obtained using a 2D Spin Echo imaging sequence with the following parameters: in-plane resolution = 0.9375 mm, slice thickness = 2.5 mm, TR = 5800 ms, TE = 96 ms, 52 slices, FOV = 240 × 240 mm, b-value = 1000 s/mm^2^. Two sets of scans with 4 b0 images and 32 diffusion sampling directions each were obtained with opposite phase-encoding directions. DWI data were then preprocessed using tools from the the FMRIB Software Library (FSL) (Jenkinson et al., 2012; Smith et al., 2004), detailed below. From these pairs of images with reversed phase-encoding blips the susceptibility-induced off-resonance field was estimated using a method similar to that described in (Andersson et al., 2003) as implemented in FSL’s topup tool. The resulting susceptibility field was used with FSL’s eddy_correct tool (Andersson and Sotiropoulos, 2016) to simultaneously correct DWI volumes for subject movements and susceptibility- and eddy current-induced distortions.

DWI data was resampled to 2 mm isotropic resolution and reconstructed with DSI Studio software (http://dsi-studio.labsolver.org/) using *q*-space diffeomorphic reconstruction (QSDR) (Yeh and Tseng, 2011). First, QSDR performs reconstruction in native space where the quantitative anisotropy (QA) for each voxel is computed. These QA values are used to warp DWI to a high angular resolution template in Montreal Neurological Institute (MNI) space using a nonlinear registration algorithm similar to that described in (Friston, 1994). QSDR was chosen over diffusion tensor-based approached because the reconstructed spin distribution functions can resolve crossing, branching and merging fiber populations (Yeh and Tseng, 2011). A deterministic fiber tracking algorithm (Yeh et al., 2013) was used with whole brain seeding with a total of 10000000 seeds, an angular of 70 degrees, step size of 1 mm and quantitative anisotropy threshold of 0.1. The fiber trajectories were smoothed by averaging the propagation direction with 10% of the previous direction. Tracks with length shorter than 10 or longer than 300 mm were discarded.

#### Functional MRI

Whole-brain blood-oxygen-level dependent (BOLD) functional images were acquired using a T2* weighted gradient-echo echo-planar imaging sequence optimized to reduced susceptibility artifacts with the following parameters: TR = 2000 ms, TE = 25 ms, flip angle = 82°, FOV = 220 × 220 mm, acquisition matrix 64 × 64, slice thickness = 3 mm, gap = 0 mm, 44 axial slices, 180 volumes per run. For anatomical localization, a high-resolution T1w structural scan was also acquired (described above). During this scan, subjects were asked to view a fixation cross on a blank background for 8 minutes. Subjects were instructed to keep their eyes open and focused on the cross, and to try not to think of anything in particular for the duration of the scan. Functional MRI (fMRI) data preprocessing and analysis were performed using the CONN Toolbox (www.nitrc.org/projects/conn) (Whitfield-Gabrieli and Nieto-Castanon, 2012), which employs procedures from the Statistical Parametric Mapping software (SPM12; Wellcome Trust Center for Neuroimaging, London, UK), using the standard preprocessing and denoising pipelines as detailed in (Nieto-Castanon, 2020) and described as follows.

1) fMRI images were first coregistered to the first volume of the series as reference using b-spline interpolation and susceptibility distortion-by-motion interactions were corrected by resampling the data to match the estimated deformation field of the reference volume (Andersson et al., 2001). 2) Slice-timing correction was performed by time-shifting and resampling data using sinc-interpolation to the middle of each acquisition time (Henson et al., 1999). 3) The effects of outlier scan-related nuisance (‘scrubbing’) covariates were identified by quantifying frame-wise displacement and global BOLD signal changes (Power et al., 2014). Images with displacement greater than 0.9 mm and signal change greater than 5 standard deviations from the mean were labeled as outliers. 4) fMRI images were then co-registered to the T1w structural imaging data using an affine tranformation as described in (Collignon et al., 1995; Studholme et al., 1998). T1w images were next registered to MNI space and segmented into grey matter, white matter and cerebrospinal fluid (CSF) tissue classes using a non-linear spatial transformation as described in (Ashburner and Friston, 2005). The resulting transformation was next used to warp the native structural registered functional volumes to MNI space. T1w and functional images were resampled to 1 mm and 2 mm isotropic voxel resolution, respectively. 5) Functional data were then smoothed using spatial convolution with a Gaussian kernel of 8 mm full width half maximum (FWHM) in order to increase BOLD signal-to-noise ratio and reduce the residual effects of inter-subject anatomic variability (Nieto-Castanon, 2020). 6) Functional data were then corrected for confounding effects by regressing out components derived from estimated subject motion parameters, outlier images (‘scrubbing’), white matter and CSF BOLD signal and estimated physiological noise using the aCompCor and tCompCor methods in the CONN toolbox (Behzadi et al., 2007; Chai et al., 2012). The mean global BOLD signal was not regressed out at it can result in artificial bias and diminution of meaningful neural signal (Murphy et al., 2009; Chai et al., 2012). 7) A temporal band-pass filter was applied to remove BOLD signal below 0.008 Hz or above 0.09 Hz in order to capture slow-frequency neural fluctuations while minimizing the presence of physiological and motion related noise in the functional data (Nieto-Castanon, 2020). Filtering was applied after regressing out confounding effects to avoid frequency mismatch in the resulting denoised data (Hallquist et al., 2013).

### Defining regions of interest

Cortical areas were defined by the HCP-MMP1.0 parcellation comprising of 360 (180 per hemisphere) regions of interest (ROIs) which localizes brain regions on on inflated brain surfaces (Glasser et al., 2016). Because the DWI reconstruction and tractography methods used in this manuscript require a parcellation in 3D coordinate space, a surface to volume projected version of the HCP-MMP1.0 atlas registered to MNI space and corrected for errors arising from surface-voxel misalignment (as implemented in DSI Studio) was used for the present analyses, as done in previous studies (Huang et al., 2021; Jitsuishi et al., 2020; Jitsuishi and Yamaguchi, 2021; Ghulam-Jelani et al., 2021; Wu et al., 2019). However, it has been shown that using volume-based HCP-MMP1.0 parcellations have higher cortical areal localization uncertainty than surface-based approaches (Coalson et al., 2018). As such, we limit the investigation of individual cortical areas to a much coarser parcellation scheme (*n* = 44 total ROIs, 22 per hemisphere) by aggregating the 360 original ROIs by their cortical region label as defined by the HCP-MMP1.0 atlas (Glasser et al., 2016). The exception to this rule is with the right area 8BM which was found to be the hub of diffusion impairment of the subnetwork discovered in this study. To verify the localization of the right area 8BM volumetric parcel, we determined the percentage of voxels that are labeled by ROIs from derived from individual subject surface-registered HCP-MMP1.0 parcellations mapped to MNI volumetric space using tools from Connectome Workbench (Glasser et al., 2013) and DSI Studio software. In doing this, we found 81.5% of the right area 8BM voxels to be correctly mapped (see Supplementary Information for details).

### Structural and functional connectivity matrices

Using the 360 ROIs from the HCP-MMP1.0 parcellation as described above, 360 × 360 matrices encoding the connectivity between each pair of brain regions were created for each subject. For DWI structural data, each connection was given by the count of reconstructed white matter tracts, normalized by the median tract length, between ROIs. Functional connectivity was defined by as the *r* statistic from pairwise Pearson correlations on the mean BOLD time series data of voxels within each ROI.

### Network notation

A brain network may be represented as a graph with nodes being grey matter regions and edges being the connection between these regions. For structural connectomes, edge weights are assigned based on the number of fiber counts between nodes, normalized by the median fiber length. For functional connectomes, edge weights are the Pearson correlation *r* of the time series of BOLD signals between nodes. Formally, a graph is defined as *G* = (*V, E*) where *V* is the set of nodes of size *N* and *E* is the set of edges linking nodes in *V*. In addition, *w* : *E* → ℝ, is a weight function that assigns weights to the *E* according to the modality of imaging from which the brain graph is constructed. *G* can then be encoded as an adjacency matrix, *A* ∈ ℝ^*NxN*^, where

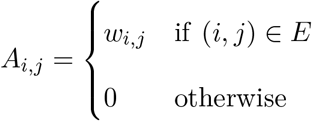

each entry *A*_*i,j*_ corresponds to the connection weight between nodes *v*_*i*_ and *v*_*j*_. The diagonal strength matrix is then defined as 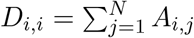 which can be used to define the graph Laplacian matrix, *L* = *D* − *A*, and, the normalized graph Laplacian is defined as 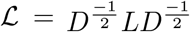. The eigendecomposition of the graph Laplacian is given by ℒ = *U* Λ*U*, where *U* is a matrix with columns as eigenvectors and Λ is the diagonal matrix of eigenvalues. The spectral properties of the normalized graph Laplacian have been extensively studied (Chung and Graham, 1997).

### Diffusion in networks

Diffusion in networks over time can be determined analytically using the heat equation given by

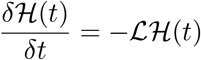

where H(*t*) is the heat kernel and fundamental solution to the heat equation, and *t* is time. The heat kernal can be understood as describing the flow of information between all nodes of a network over time, taking into account all possible pathways of information flow into and out of all nodes in the network. Throughout this manuscript, heat or information with be used interchangeably in this context. The heat kernel is then given by

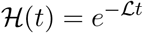

which can then be used to solve for the heat distribution on network nodes, *h*(*t*) after time = *t*, given an initial condition *h*(0):

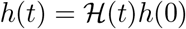

The product of an element of the heat kernel and initial condition vector, ℋ(*t*)_*i,j*_ ∗ *h*(0)_*i*_, then represents the amount of heat at the *j*^*th*^ node at time = *t* that has diffused from the *i*^*th*^ node. The heat kernel can be computed as a sum of the outer product of eigenvectors of the graph Laplacian, scaled by exponentiating the corresponding eigenvalues with time:

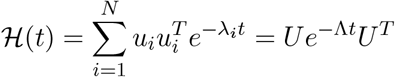

### Structure to function mapping

In this study, we use the methods proposed by Abdelnour and colleagues (Abdelnour et al., 2014) (Abdelnour et al., 2018) to identify the graph diffusion-based mapping of functional connectivity from the structural basis. Briefly, the observed functional activity in rs-fMRI, i.e., the transfer of information from brain region (node) *i* to region *j* as measured by pairwise correlation of temporal BOLD signals, can modeled by first order diffusion-like dynamics given by:

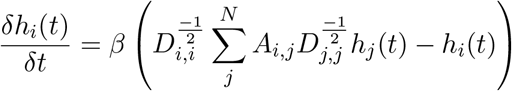

where *β* is a diffusion constant. This extends to the entire network then as the heat equation,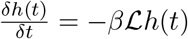. The mapping from the structural diffusion dynamics, yielding the estimated functional connectome, 𝒞_*est*_,is then given by (Abdelnour et al., 2014)

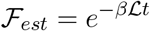

and has been updated as (Abdelnour et al., 2018)

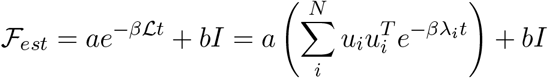

where *a* and *b* are additional model parameters and *I* is the identity matrix. Note that the summation allows for the exclusion of eigenvector-eigenvalue pairs. As in (Abdelnour et al., 2018), we leave out the first eigenvector with corresponding zero valued eigenvalue, as it largely represents uniform background connectivity (captured by *b* parameter above) and is typically regressed out of rs-fMRI data. The model is then fit by minimizing the normalized predictive error with respect to the model parameters given by:

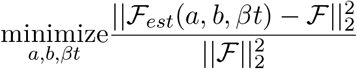

where ℱ is the empirical rs-fMRI connectivity matrix. This model is fit for each individual to obtain subject specific *βt* parameters, used in the diffusion-based embedding discussed below.

### Diffusion-based network embedding

The embedding of nodes based on the diffusion properties of a graph has been studied extensively for dimensionality reduction and clustering of multi-dimensional data (Belkin and Niyogi, 2003; Ng et al., 2002; Luo et al., 2003). These approaches center around the embedding of network vertices via the eigenvectors of the graph Laplacian. Each element of an eigenvector corresponds to the coordinate of the corresponding node such that nodes that are closer together by geodesic distance on the underlying graph or manifold have more similar coordinate values, i.e., are closer together in Euclidean distance, in the embedding. A subset of these eigenvectors of size *k* = 1, 2, …, *N* can then be used to embed nodes in ℝ^*k*^. In the context of this manuscript, then, brain nodes that are able to more efficiently pass information between one another are embedded closer together via the Laplacian eigenmodes of a brain network (Wang et al., 2017).

To compute the temporally dependent diffusion-based embedding for a network, we follow the methods discussed by Xiao and colleagues, where node coordinates are obtained from the Young-Householder decomposition of the heat kernel (Xiao et al., 2010, 2005). The embedding matrix, *Y* = (*y*_1_|*y*_2_|…|*Y*_*N*_), with columns as embedding coordinate vectors for network nodes can determined with the heat kernel by ℋ(*t*) = *Y* ^*T*^ *Y*. The Euclidean distance between nodes *i* and *j* is then

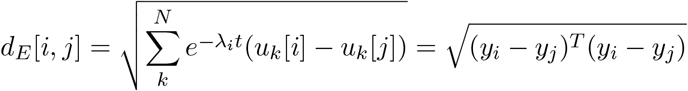

where the distance in each dimension is scaled by exponentiating the product of the corresponding eigenvalue and diffusion time, *t*. A pairwise Euclidean distance matrix, 𝒟, of embedded nodes is computed for each subject with the time parameter *βt* obtained via the structure to function mapping as described above. This provides a newly defined structural connectome where edges are distances in the embedding space spanned by the eigenvectors of the graph Laplacian, and the scale of distance in each embedding dimension is dependent on the time parameter, *βt*, for each subject:

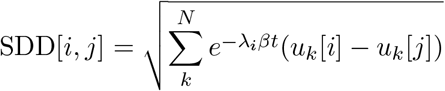

### Subnetwork identification

The structural diffusion distance (SDD) connectome edges then represent the distance between brain regions in diffusion embedding at the diffusion depth that best represents empirical resting-state functional connectivity. Many previous findings from studies of IPs implicate perturbations in resting state functional connectivity, especially within the DMN, which is defined by activity at rest. As such, SDD connectomes then used for further analysis to identify structural subnetworks with aberrant diffusion characteristics pertinent to rs-fMRI dynamics using the Network-based Statistic (NBS) algorithm (Zalesky et al., 2010). Briefly, NBS is carried out by computing a t-statistic at each network edge, applying a pre-determined threshold to the resulting t-statistics and determining the connected components formed by supra-threshold edges. To determine the significance of each identified subnetwork, the size of each component is compared to a null distribution of maximum sized connected components obtained by shuffling group labels for t-statistic calculation. NBS was carried out using SDD connectomes in HC versus PT at baseline, using a left sided t-test with t-statistic thresholds of *t* = {3.0, 3.5, 4.0, 4.5}. In doing this, a subnetwork, *S* ⊂ *G*, where *G* is the full brain network graph, with significantly greater embedding distances, or significantly decreased diffusion capacity, in PT relative to HC was.

#### NBS subnetwork hubs

Hub node identification for brain regions within the significant NBS subnetwork was performed using edges from both the SDD and standard fiber count structural connectomes, and hub or centrality metric values were then averaged by group. For SDD connectomes, strength, or weighted degree, str_*SDD*_(*n*), for the *i*^*th*^ node, *n*_*i*_, in *S* is defined as the sum of all edge weights (distances) that are within the identified subnetwork, *S*, i.e.,

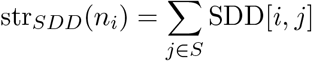

A high strength value would then indicate that a node has relatively less diffusion between other nodes in the subnetwork, thus identifying nodes that may have the greatest diffusion impairment. For structural connectomes, standard measures of nodal strength, betweenness centrality, local efficiency and clustering coefficient (Bullmore and Sporns, 2009; Sporns, 2018) were computed on the whole brain network graph and subgraph, *S* ⊂ *G*, for each node in *S*. For each of the above scenarios, metrics were also determined by cortical region by averaging region of interest (ROI) level metrics by their cortex assignment as defined by the HCP-MMP1.0 360-node parcellation (Glasser et al., 2016).

### Heat kernel edge-based analyses

In order to study more granular characteristics of subnetwork diffusion, heat kernel values between nodes within the significant NBS subnetwork were investigated using t-tests in HC vs PT at baseline. Correlates of baseline symptom severity (using PT only) of and treatment response (defined as 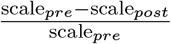 were found by computing non-parametric Spearmen rho statistics between symptom scales and heat kernel values. As above, where cortex-averaged metrics were computed, heat kernel edge-based analyses were conducted cortex-averaged heat kernels in order to simplify interpretation and reduce the number of mass univariate tests performed. To further focus this analysis, statistics other than baseline t-statistics were only computed on heat kernel values corresponding to pairwise links between SSD hubs (as determined above) and all other subnetwork nodes. P-values for all statistics were corrected for multiple comparisons using False discovery rate correction (FDR) (Benjamini and Hochberg, 1995).

### Subnetwork targets for supplemental heat

To identify potential brain regions for neuromodulatory treatment, we model a brain subnetwork receiving supplemental heat by simply adding a heat kernel modifying matrix, whose rows are made by repeating rows of the original heat kernel, to the original heat kernel. As discussed previously, the product of an entry of the heat kernel matrix, ℋ(*t*)[*i, j*], and the heat at node *i* at time *t* = 0, *h*(0)_*i*_, indicates the amount of heat transferred from node *i* to node *j* at time *t*. The product of the *i*^*th*^ row of the heat kernel matrix, ℋ(*t*)[*i*, :], and *h*(0)_*i*_ then yields the distribution of heat values for each *j*^*th*^ node in the network. If the *i*^*th*^ node of a network is supplied with supplemental heat independent of the initial distribution of heat on the network, *h*(0), the network heat distribution at time *t* is given by

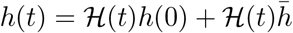

where 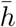 is a vector of all zeros except for the *i*^*th*^ whose value is the amount of supplemental heat at node *i*. Using this model, we can then identify both a node and supplemental heat value that may best ‘normalize’ a heat kernel representing impaired diffusion processes to a reference heat kernel. In this study, we use the mean HC heat kernel matrix as the reference and identify patient-specific nodes and supplemental heat values for optimal correction of the diffusion dynamics encoded in the heat kernel, described as follows.

Given the mean HC heat kernel, ℋ_*HC*_, and the *k*^*th*^ patient’s heat kernel, 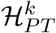, we compute the residual heat kernel matrix as 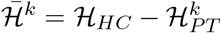. For the *i*^*th*^ node in the subnetwork, we construct the heat kernel modifier matrix as 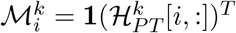 as the outer product of the all ones vector and the *i*^*th*^ row of the *k*^*th*^ patient’s heat kernel matrix. In order to find the optimal supplementary heat value, 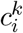 for a given node and subject, we minimize the following objective function.

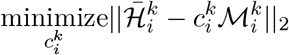

Which can be easily solved analytically by

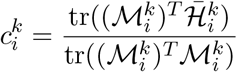

where tr(·) denotes the matrix trace.

It is important to note, that the above model is also constrained such that only positive values in the residual matrix are used for optimization, as this simple network diffusion model only allows for the addition of heat.

This process is repeated for each unique node-patient pair, and the mean or patient-specific optimal node and corresponding supplemental heat value can be determined. We have experimentally observed that the minimum error between residual and heat kernel modifier matrices has very little variation between brain regions, and we define optimal region as the node with the lowest value supplemental heat value, such that neuromodulatory efficacy could be achieved with lower amounts of stimulation and thus potentially yield fewer undesired effects.

#### Visualizing effects of heat supplementation

To assess whether the significant group differences at baseline between HC and PT heat kernel values are corrected with heat supplementation, we add the product of the optimal supplemental heat value, 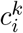, at the *i*^*th*^ node, and the heat kernel modifier matrix, 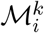 to each *k*^*th*^ patient’s heat kernel. We then compute a t-statistic for each heat kernel entry, as discussed above.

## RESULTS

Data analyzed in this study are taken from a previously conducted Research Domain Criteria (RDoC) clinical trial on predictors of IP treatment outcomes (ClinicalTrials.gov identifier: NCT01903447) in which PT were randomized to either 12 weeks cognitive behavioral therapy (CBT) or selective serotonin reuptake inhibitor (SSRI) treatment. In addition, the patient population has heterogeneous clinical presentations and IP comorbidity in order to study common pathological features without regard to traditional diagnostic categories. Baseline rs-fMRI and DWI scans as well as IDAS-II Panic and Depression scores were assessed from both HC (*n* = 22) and PT (*n* = 65) subjects. The subset of PT (*n* = 50 total; *n* = 28 CBT cohort; *n* = 22 SSRI cohort) completed the IDAS-II self-reports following treatment are used for identifying correlates of treatment response. Of note, there were no group differences in age or sex between HC and PT. As expected, baseline IDAS-II Panic and Depression scores were significantly different between groups at baseline (table 1).

**Table 1:**
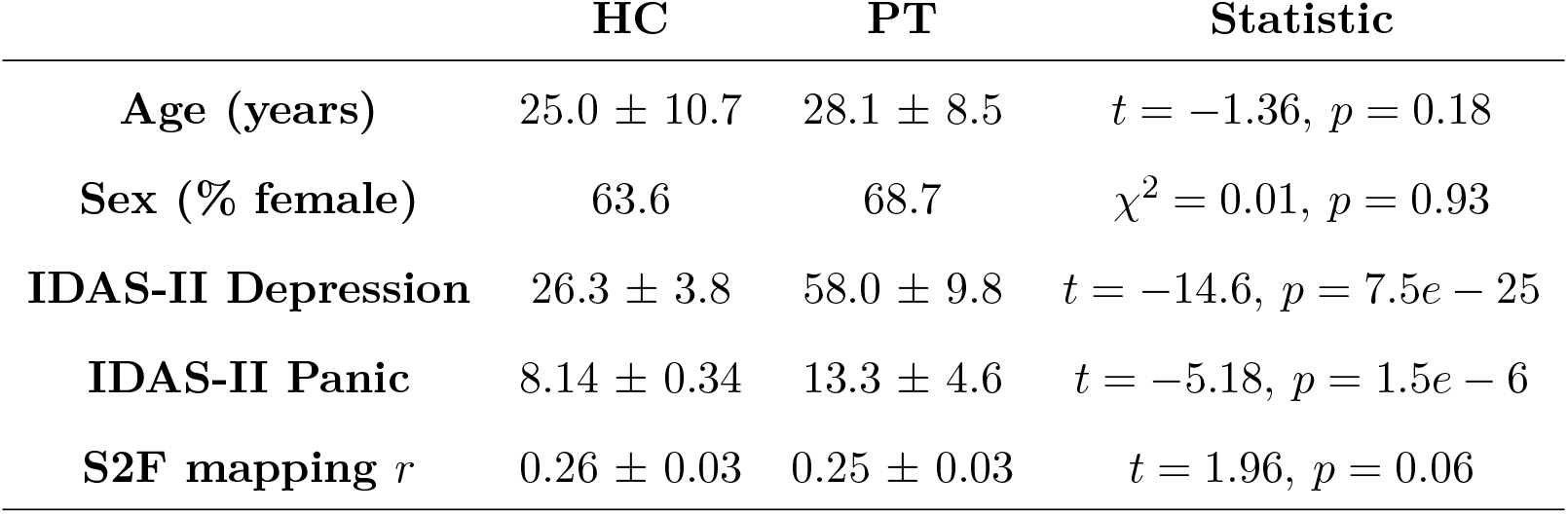
Baseline mean demographics, IDAS-II scales and structure to function mapping (S2F) Pearson *r* values by split healthy controls (HC) and patients (PT).

|  | HC | PT | Statistic |
| --- | --- | --- | --- |
| **Age (years)** | $25.0 \pm 10.7$ | $28.1 \pm 8.5$ | $t = -1.36, p = 0.18$ |
| **Sex (% female)** | 63.6 | 68.7 | $\chi^2 = 0.01, p = 0.93$ |
| **IDAS-II Depression** | $26.3 \pm 3.8$ | $58.0 \pm 9.8$ | $t = -14.6, p = 7.5e - 25$ |
| **IDAS-II Panic** | $8.14 \pm 0.34$ | $13.3 \pm 4.6$ | $t = -5.18, p = 1.5e - 6$ |
| **S2F mapping $r$** | $0.26 \pm 0.03$ | $0.25 \pm 0.03$ | $t = 1.96, p = 0.06$ |

Figure 1 provides a brief visual overview of the analytic framework for computing the diffusion-based embeddings and subsequent structural diffusion distance (SDD) connectomes used in this study, discussed in detail in the Methods section. We first compute the structure to function mapping as described in (Abdelnour et al., 2018) for each subject. The fit of the empirical rs-fMRI connectome to the rs-fMRI connectome estimated from the DWI structural connectome Laplacian eigenmodes is then determined by computing the Pearson *r* statistic between these adjacency matrices. The mean fit values for HC were slightly higher than PT (*r* = 0.26 ± 0.03 and *r* = 0.25 ± 0.03, respectively, table 1) but the difference between their mean values was not statistically significance via t-test (*p* = 0.06), indicating that the relation between resting state functional connectivity and the structural diffusion basis is likely preserved at the global scale. Finally, the distance matrix connectomes used for subnetwork discovery are computed from the structural diffusion basis at the diffusion depth, *βt*, that optimally maps structural to functional connectivity.

**Figure 1:**
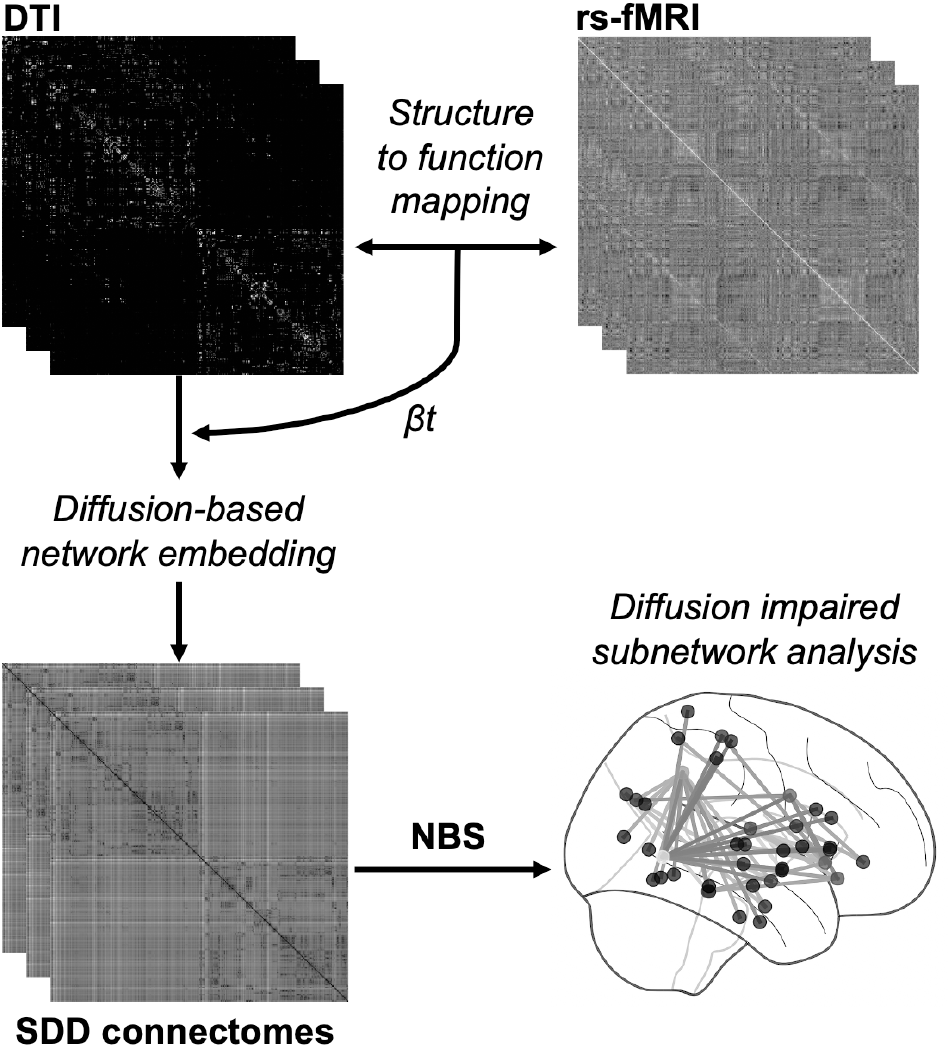
Visual overview of methodological pipeline for creating structural diffusion distance (SDD) connectomes using subject-specific optimal diffusion depths (*βt*) from the structure-to-function mapping as in (Abdelnour et al., 2018) using subject DWI and rs-fMRI connectomes, respectively. NBS is then applied to SDD connectomes for identification of subnetworks with aberrant diffusion properties for subsequent analyses. See Methods section for details.

### Identification of Diffusion Impaired Subnetwork

After SDD connectomes were computed, the Network-Based Statistic (NBS) algorithm (Zalesky et al., 2010) was used to discover subnetworks with altered diffusion properties in PT relative to HC. NBS was applied with one-sided t-tests used for edge comparison, over a range thresholds *t* = {3.0, 3.5, 4.0, 4.5}. For a t-test contrast (left-sided t-test) of HC < PT, the range of subnetwork sizes varied greatly as a function of t-statistic threshold, resulting in subnetworks from 314 to 2 nodes with thresholds *t* = 3 and *t* = 4.5, respectively. With the threshold of *t* = 4 a single significant subnetwork (SN1) consisting of 48 nodes and 73 edges was identified (Figure 2A) and was chosen for subsequent analyses because of the biological interpretability of its size, and for adequate estimated statistical power, 1 − *β* = 0.81, with a t-statistic threshold of *t* = 4. Note that because NBS was conducted on SDD connectomes, edges are pairwise distance between nodes in the diffusion-based network embedding. This indicates that SN1 represents a subnetwork whose nodes are significantly further apart in this diffusion space, i.e., diffusion of information occurs more slowly between the nodes of SN1 along the edges of SN1 in PT relative to HC. For NBS with a t-test contrast defined as HC > PT, no significant subnetworks were found.

**Figure 2:**
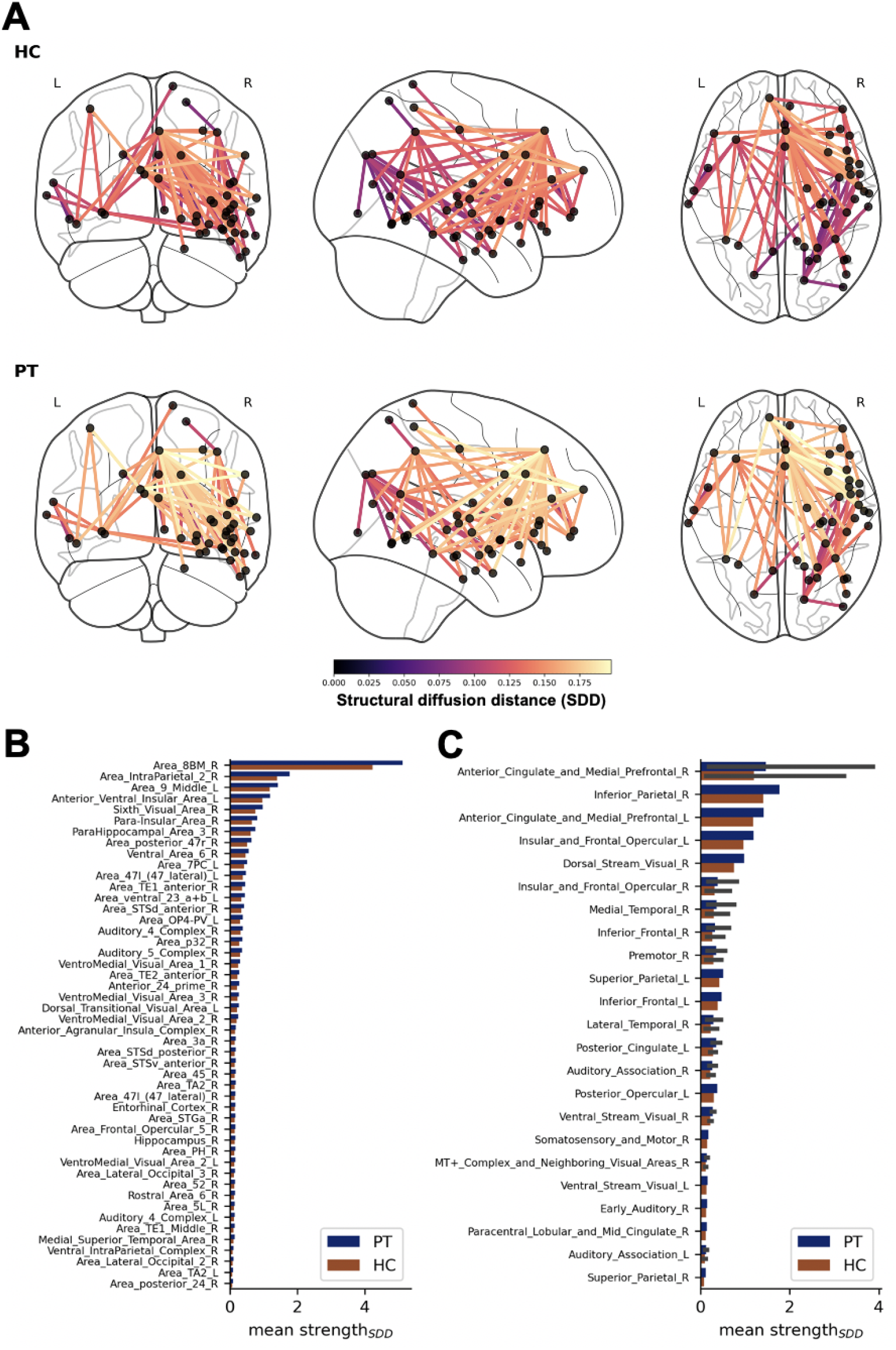
Visualizations of the significant subnetwork (SN1) found via NBS with a t-statistic threshold of *t* = 4 and contrast of HC < PT, where information diffusion is impaired in PT relative to HC. **A** Wire and ball plots of SN1 subgraphs of mean SDD connectomes in HC (top row) and PT (bottom row). Nodes indicate brain regions of SN1, and edges are colored by mean distance in from SDD connectomes (lighter/more yellow indicates greater distance). **B & C** Bar plots of mean strength (str_*SDD*_) of SN1 nodes **B** and nodes averaged by cortical region **C**, as determined by cortex assignment in the HCP-MMP1.0 parcellation (Glasser et al., 2016), calculated using edges within SN1 and the corresponding edge values of mean SDD connectomes HC (orange bars) and PT (blue bars). A greater strength indicates a greater diffusion distance from other nodes in SN1.

To determine the brain regions that are most central to the diffusion impairment in SN1, we calculated the mean strength, str_*SDD*_, of each node using the SDD values of edges in SN1 (see Methods section for details). The nodes with the greatest str_*SDD*_ values are from brain regions found in the bilateral anterior cingulate (ACC) and medial prefrontal (mPFC), right inferior parietal and left insular and frontal opercular cortices (Figure 2A). Specifically, the brain region with the greatest average str_*SDD*_ value in both HC and PT is the right area 8BM, which has recently been reported to be a core region of a ‘multiple demand’ subnetwork that is active during a broad spectrum of tasks and may play an important role in general cognitive control (Assem et al., 2020). Additionally, we found that the hubness of area 8BM was not present in structural connectomes, determined by computing graph theoretical metrics, indicating that the centrality of this brain region is unique to the SDD representations of brain networks (Figure S1).

### Baseline Heat Kernel Correlates of Symptom Severity and Treatment Response

To investigate the association of information diffusion between brain regions of SN1 and clinical scales, we compute heat kernel matrices which encode the amount of possible heat transferred between each node after a specified time scale, given by each subject’s diffusion depth, *βt*. Each value, *i, j*, of a heat kernel matrix then describes pairwise diffusion of information from the *i*^*th*^ to the *j*^*th*^ brain network node at the time scale best associated with functional connectivity. Note that heat kernels are symmetric matrices (i.e., diffusion from node *i* to node *j* is equivalent to diffusion from node *j* to node *i*). Therefore, when discussing diffusion between brain regions, the order in which they are listen is arbitrary, as network diffusion approaches model undirected diffusion within a brain network. We took a hypothesis-driven approach by focusing our correlation analyses on the heat transfer between the bilateral ACC and all other SN1 cortical regions, as the nodes in these cortical regions are central to the diffusion impairment of SN1. The heat kernel values (HKVs) are averaged by cortical region and are then used for computing Spearman correlations with IDAS-II Depression and Panic subscales, which map to distress and fear domains, respectively, based on a previously used approach to broadly divide and assess symptom domains of IPs (Figure 3) (Radoman et al., 2019; Ofrat and Krueger, 2012). In addition, we use only PT (*n* = 65) for these analyses, as mean IDAS-II subscale values expectedly differ highly significantly between HC and PT, and correlations would likely be due to group differences. Finally, we report at most the top three most significant correlations that survive FDR correction (*n* = 44 comparisons) in this section of the manuscript, but all significant correlations are available in Table S1.

**Figure 3:**
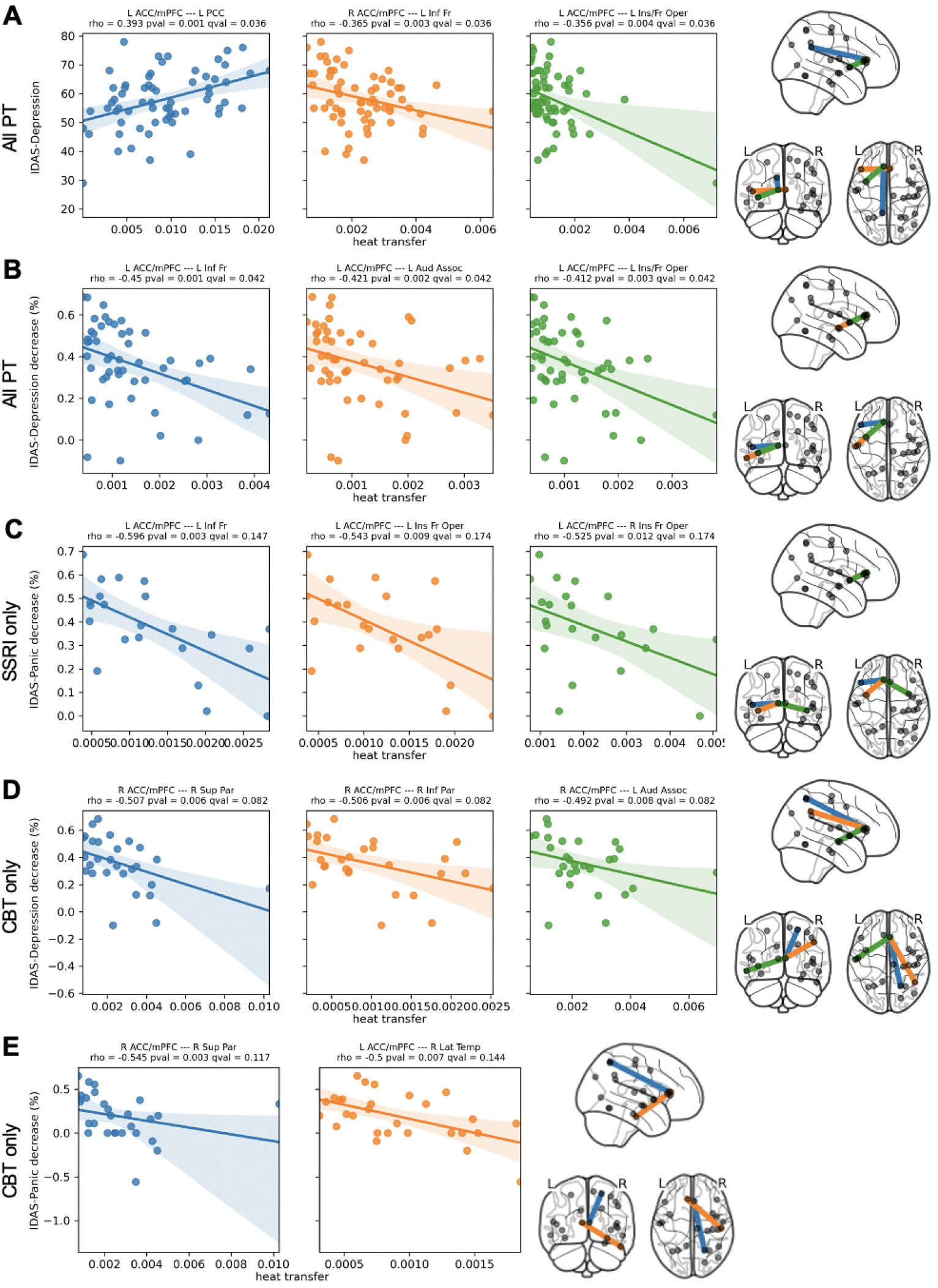
Correlation of baseline heat transfer at subject diffusion depths *βt* with pre-treatment and percentage improvement post-treatment IDAS-II subscales. The brain network figure at the end of each row shows the corresponding pair of brain for each respective correlation in the row, indicated by the color of the edge connecting the regions and the correlation plot of the same color. Cortical regions shown in the visualization are plotted according to the average position of the individual ROIs that they consist of, as per the HCP-MMP1.0 parcellation (Glasser et al., 2016). Correlations of heat transfer with: **A** pre-treatment IDAS-II Depression in PT; **B** post-treatment IDAS-II Depression percent improvement in all PT; **C** post-treatment IDAS-II Panic percent improvement in SSRI cohort only; **D** post-treatment IDAS-II Depression percent improvement in CBT cohort only; **E** post-treatment IDAS-II Depression percent improvement in CBT cohort only.

First, we determine the HKV correlates of baseline IDAS-II subscales (Figure 3A). Heat transfer between the left ACC/mPFC and insula/frontal opercula (*ρ* = −0.37, *p* = 0.003, *q* = 0.04) and the right ACC/mPFC and left inferior frontal cortex (*ρ* = −0.36, *p* = 0.004, *q* = 0.04) was negatively correlated with IDAS-II Depression scales, indicating that less information diffusion between these brain regions is associated with higher IDAS-II Depression values. A positive correlation was found with heat transfer between the left ACC/mPFC and posterior cingulate cortex (*ρ* = 0.39, *p* = 0.001, *q* = 0.04). No significant correlations were found using IDAS-II Panic.

To determine whether baseline HKVs could predict response to treatment, we determined correlations between heat transfer and percent improvement in IDAS-II subscales in the subset of PT (*n* = 50) that completes post-treatment IDAS-II self-reports (calculated as 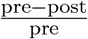, where pre and post are the IDAS-II subscales before and after 12 weeks of treatment, respectively). We first grouped both SSRI and CBT therapy cohorts of PT together to study common correlates of treatment response. Significant negative correlations with IDAS-II Depression and heat transfer between the left ACC/mPFC and inferior frontal cortex (*ρ* = −0.45, *p* = 0.001, *q* = 0.04), left auditory association cortex (*ρ* = −0.42, *p* = 0.002, *q* = 0.04) and insula/frontal opercula (*ρ* = −0.41, *p* = 0.003, *q* = 0.04) were found (Figure 3B). Similar to baseline symptom correlations, no significant associations were found using IDAS-II Panic.

Next, we segregated PT by cohort to investigate treatment specific HKV predictors of therapeutic response. Interestingly, in the SSRI cohort (*n* = 22), significant negative correlations were found only with IDAS-II Panic improvement and heat transfer between the left ACC/mPFC and inferior frontal cortex (*ρ* = −0.60, *p* = 0.003, *q* = 0.15), insula/frontal opercula (*ρ* = −0.54, *p* = 0.009, *q* = 0.17) and right insula/frontal opercula (*ρ* = −0.53, *p* = 0.012, *q* = 0.17) (Figure 3C). In the CBT cohort (*n* = 28), we observed significant negative associations with IDAS-II Depression percent improvement and heat transfer between the right ACC/mPFC and superior parietal (*ρ* = −0.51, *p* = 0.006, *q* = 0.09), inferior parietal (*ρ* = −0.51, *p* = 0.006, *q* = 0.09) and left auditory association (*ρ* = −0.50, *p* = 0.008, *q* = 0.09) cortices (Figure 3D). We also found significant negative correlations with IDAS-II Panic percent improvement and heat transfer between the right ACC/mPFC and superior parietal cortex (*ρ* = −0.55, *p* = 0.003, *q* = 0.12) and the left ACC/mPFC and right lateral temporal cortex (*ρ* = −0.50, *p* = 0.007, *q* = 0.14) (Figure 3E). Interestingly, the brain regions involved significant correlations of treatment response in all PT and the SSRI cohort are all Frontal cortical regions that have been tied to emotion regulation in MDD (Roberts et al., 2017; Rolls et al., 2020; Helm et al., 2018), while the regions found with the CBT cohort are members of canonical RSNs. For example, the inferior parietal and lateral temporal cortices are members of the default mode network (DMN), while the superior parietal cortex is part of the dorsal affective network (Dutta et al., 2014; Fedota and Stein, 2015). Please note that we increased the threshold for significance after FDR correction from *q <* 0.05 as for baseline and percentage improvement of IDAS-II subscales in all PT to *q <* 0.2 given the decreased sample size and decreased power of these correlations. As such, these results should be understood as exploratory and preliminary.

### Identification of Subnetwork Targets for Modulation

Once we had characterized the disrupted information diffusion of SN1 in IP PTs, we next sought to determine brain subnetwork regions that may serve as neuromodulatory targets to normalize aberrant subnetwork information diffusion properties towards those found in HC. To this end, we start by computing the difference between the mean HC heat kernel matrix and each *k*^*th*^ PT’s heat kernel matrix to obtain a residual heat kernel matrix. A favorable modulation strategy would then add supplemental diffusion activity to a PT’s heat kernel with a pattern that closely resembles the deficits encoded in a PT’s residual matrix. To identify such a strategy, the error of the difference between this residual matrix and the product of a heat value, 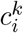, and a heat kernel modifier matrix, which encodes the effect of supplemental heat at a the *i*^*th*^node on the network, is minimized over 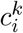 and repeated for all possible target nodes in order to identify an optimal brain region and heat value for stimulation. Once a modulation strategy is determined, the product of the modifier matrix and the heat value are then added to a PT’s heat kernel to model the effects of the intervention (see Methods section for details). For each potential target brain region, we then obtain a heat and error value corresponding to the optimal effectiveness of a brain region as a modulation target. As the error values for all target node tested were empirically observed to be uniform in distribution with small regional variance (Figure 4C), we defined optimality as the brain region requiring the smallest amount of supplemental heat. Furthermore, this allows us to identify brain region neuromodulatory targets that potentially more efficiently disseminate stimulation with fewer off-target effects from a greater modulatory energy.

**Figure 4:**
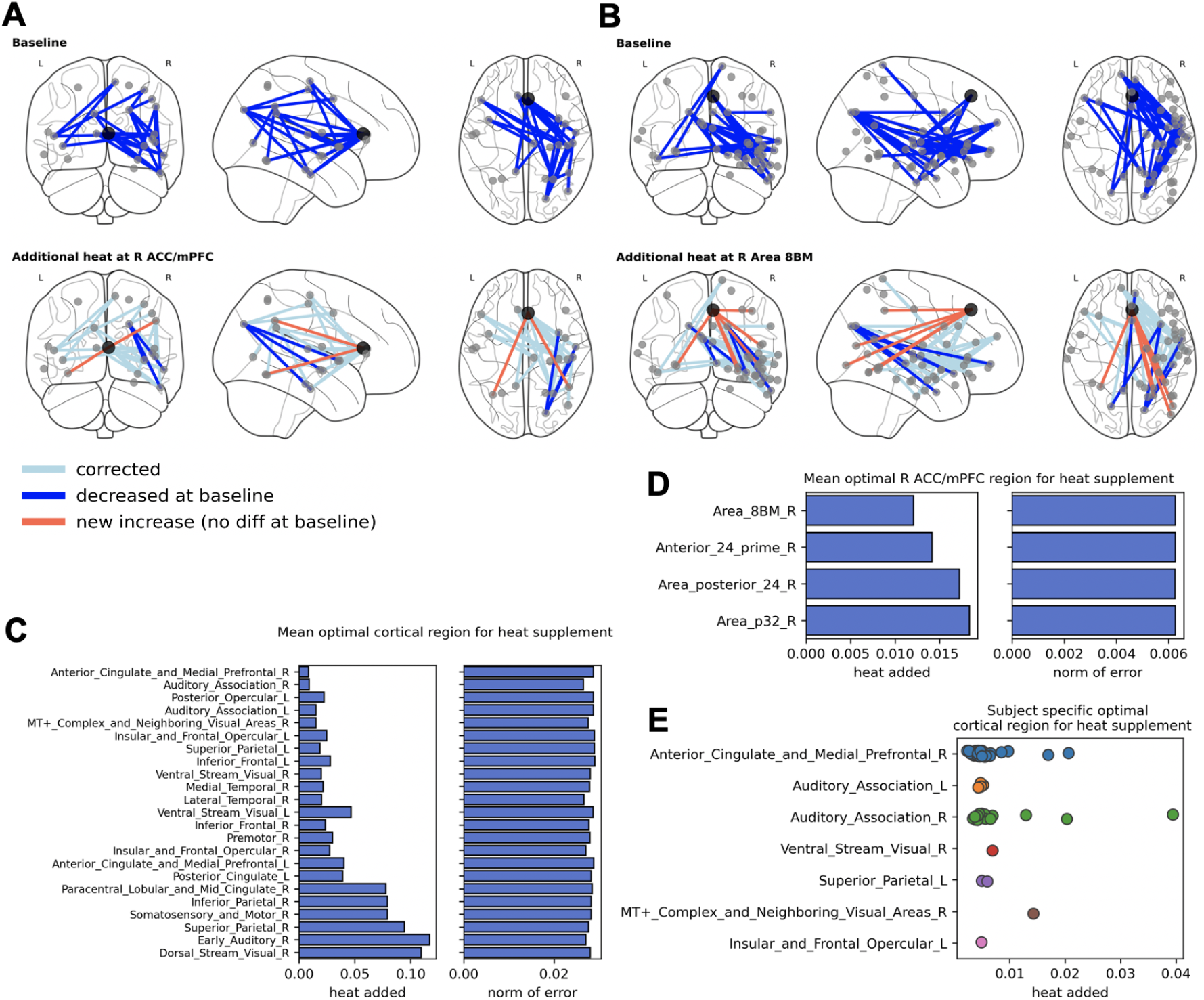
Overview of the identification of SN1 brain region targets for heat-based modulation and subsequent correction of information diffusion deficits. **A & B** Connectome plot visualizations of baseline (first row) and post-heat supplementation (second row) significant differences in heat kernel values with **A** right ACC/mPFC or **B** right area 8BM as modulation targets. Nodes are brain regions of SN1 and edges connecting them are present if a significant difference (defined as t-test p-values that survive FDR correction with *q <* 0.05) exists between HC and PT corresponding heat kernel values. Larger/darker node is the target for modulation. Edge colors indicate the type of significance (dark blue: present at baseline (first row) or remains after modulation (second row) (HC > PT); light blue: present at baseline but corrected following modulation; orange: newly significant following modulation (PT > HC)). **C & D** barplots for mean PT heat (left) and norm of error (right) values for C all mean cortical regions or D subregions of the right ACC/mPFC in SN1. **E** Scatter plot indicating subject-specific optimal modulation target heat values grouped by brain mean cortical regions in SN1.

We first conducted this analysis on the mean cortical regions of SN1 as target nodes and found the right ACC/mPFC, which was identified as a hub of SN1 by mean str_*SDD*_, to be the optimal region to target for heat supplementation, with the lowest mean heat across all PT (Figure 4A and C). In addition, we determined subject-specific optimal brain regions and the right ACC/mPFC was also found to be the modal optimal target (34 out of 65 PT) (Figure 4E). Next, we applied the same target node identification procedure to the individual brain regions of the right ACC/mPFC in SN1. We found that the right 8BM was the optimal nodal modulatory target (Figure 4B and D). Note that the right 8BM was an SN1 hub with the greatest str_*SDD*_ of all individual subnetwork nodes.

### Correction of Subnetwork Diffusion Impairment

As a final step in our analysis, we sought to confirm whether the heat modulation approach identified as above successfully normalizes subnetwork information diffusion in PT to that of HC statistically. To carry this out, we performed left sided t-tests (contrast: HC > PT) at each heat kernel value (HKV) between the brain regions of SN1 at baseline in HC versus PT. Only this direction of t-test was assessed at baseline as this modulation model can only add heat, and is not presently able to correct hyperconnected HKVs in PT. We then modified PT heat kernels at the optimal target nodes and heat values as described above and repeated the edgewise t-test analysis with left and right sided t-tests in order identify significantly increased HKVs in PT relative to HC as a result of heat supplementation. An HKV was defined as significant if it survived FDR correction (*q <* 0.05) with 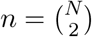comparisons, where *N* is the number of brain regions in the subnetwork.

We conducted this analysis first on mean cortical regions using the right ACC/mPFC (*N* = 23) as a target for modulation and identified 24 significant baseline HKV differences (Table S2). Following heat supplementation, 19 of the original 24 HKV differences were no longer significant (Figure 4A, Table S2). Three HKVs remained significant in the same direction as baseline: connections between the right dorsal visual stream and the right auditory association area, insula/frontal opercula and lateral temporal cortex. On the other hand, 2 HKVs gained new significance (PT > HC) following modulation; connections between the right ACC/mPFC (target) and the left ventral visual stream and the right inferior parietal cortex.

We next conducted the same analysis on individual brain regions in SN1 (*N* = 48), using the right area 8BM as the modulatory target (Figure 4B, Table S3). Of the 31 HKVs differences at baseline, 26 were corrected following heat modulation. Five HKVs remained significant following modulation, which predominantly follow the regional pattern as described above. All 6 of the newly significant (PT > HC) HKVs involved connections with the right area 8BM (target node).

## DISCUSSION

This research provides preliminary evidence of a transdiagnostic subnetwork deficit, that resembles the cingulo-opercular network, characterized by information diffusion impairment of the bilateral ACC and mPFC. Central to this impairment is more specifically the right area 8BM, a key brain region involved in organizing a broad spectrum of cognitive tasks, which may underlie previously reported dysfunction of multiple brain circuits in the IPs. In addition, this is also the first report of using network diffusion models to study psychiatric disorders. We also demonstrate that models of neuromodulation involving targeting this brain region normalize PT diffusion dynamics towards those of healthy controls. These analyses provide a framework for multimodal methods that identify diffusion disrupted subnetworks and potential targets for neuromodulatory intervention based on previously well-characterized methodology.

From our analyses we discovered a subnetwork (SN1) with increased diffusion embedding distance, i.e., decreased information diffusion capacity at the time scale required for optimally capturing functional connectivity patterns, in HC relative to IP PT at baseline. Hub regions (as defined by HC or PT mean str_*SDD*_, indicating regions with the greatest total diffusion impairment) of SN1 include the bilateral ACC/mPFC and insula/frontal opercula, which bears resemblance to the cortical aspect of the cingulo-opercular network (CON) (Figure 2). The CON, which includes the dorsal ACC, anterior insula, PFC, hypothalamus, thalamus and amygdala has been shown to contribute to many brain functions, including the processing of pain and negative affect, as well as the maintaining general cognitive control during goal-oriented behaviors (Dosenbach et al., 2007). Dysfunction of the CON has also been demonstrated in IPs, including MDD (Hamilton et al., 2013) and anxiety disorders (Sylvester et al., 2012). The dorsal ACC, a core hub of the CON, has been shown to be critical in both MDD (Wu et al., 2016) and anxiety disorders (Sylvester et al., 2012). In particular, investigations of emotion regulation in anxiety disorders have demonstrated that increased ACC activity (Burkhouse et al., 2018b) and coupling to anterior insula activity (Klumpp et al., 2012, 2013b) is present in HC relative to PT, indicating a potential regulatory role for the ACC during cognitive control of emotion processing within the CON.

SN1 also includes brain regions that are part of other functional brain networks, such as the lateral temporal and inferior parietal cortices (a hub of SN1), part of the DMN, and the superior parietal cortex, part of the dorsal affective network, in addition to the inferior frontal cortex, which is important for the coordination of executive function and emotion processing (Seeley et al., 2007; Roberts et al., 2017). Of note, the inferior parietal cortex is also a member of the frontal parietal network, an executive function network that antagonistically deactivates the DMN (Barkhof et al., 2014). Increased activity of the DMN, a network defined by functional connectivity at rest associated with both rumination and worry, relative to other RSNs has been implicated in MDD (Dutta et al., 2014) and anxiety disorders (Kim and Yoon, 2018). Studies of MDD have also indicated the widespread disruption in the salience and central executive networks (Kaiser et al., 2015; Gong and He, 2015). Indeed, accumulating evidence suggests that dysfunctional coordination between multiple brain networks may better explain the pathophysiology of IPs.

Our findings from interrogating interregional information transfer within SN1 indicate that connections between the bilateral ACC/mPFC and brain regions involved with canonical brain networks are associated with baseline symptom severity. IDAS-II Depression scores correlated positively with heat transfer between the left ACC/mPFC and the posterior cingulate. Reduced connectivity between the posterior cingulate, a DMN hub, and DLPFC (Leech and Sharp, 2014) and other frontal regions (Yang et al., 2016) has been previously reported in MDD. In the macaque, retrograde tracing studies have revealed afferent connections to the 8BM subregions of the ACC/mPFC from the posterior cingulate (Eradath et al., 2015). Taken together, our results may indicate that depressive symptoms in the IPs at least partly result from the hyperconnectivity of the DMN to the MDN and CON, which may skew cognitive resources away from executive control of emotion processing. On the other hand, negative correlations were found with IDAS-II Depression and heat transfer between ACC/mPFC and the inferior frontal cortex and insula/frontal opercula. These results are similar to those from previous studies, where the ACC activity was found to negatively correlate with depression symptom scales (Wu et al., 2016) and with anxiety symptom scales in the context of its potentially antagonistic role with the insula in the CON (Klumpp et al., 2012, 2013b). Overall, these baseline associations provide evidence for ACC/mPFC activity and connectivity to regions important for emotion regulation as critical to transdiagnostic depression symptoms in the IPs.

Our results also indicate that internodal information transfer of the bilateral ACC/mPFC in SN1 is associated with therapeutic response in both treatment modality agnostic and specific manners. Interestingly, predictors of general and SSRI specific treatment response involved similar brain regions to those associated with baseline symptom severity. In all PT, negative correlations were found with IDAS-II Depression symptom improvement and the heat transfer between the left ACC/mPFC and the inferior frontal cortex, insula/frontal opercula and auditory association cortex. As with all PT at baseline, no associations were found with IDAS-II Panic. In the SSRI cohort, however, significant correlations were only found using IDAS-II Panic improvement, yielding negative associations with connections between similar regions as with all PT: heat transfer between the left ACC/mPFC and the inferior frontal cortex and bilateral insula/frontal opercula. These findings indicate that lower baseline information transfer between the ACC/mPFC and CON or inferior frontal cortex are predictive of general and SSRI specific responsiveness to treatment, and these connections may be the substrate of treatment action. Interestingly, associations of baseline internodal information transfer and treatment response in the CBT cohort were found with unique brain regions compared to all PT and the SSRI cohort. Significant associations were also found using both IDAS-II Depression and Panic scales. Depression improvement was negatively associated with information transfer between the right ACC/mPFC and superior and inferior parietal cortices, regions of the dorsal affective network and DMN. Using IDAS-II Panic improvement, we found significant negative correlations with information transfer between the right ACC/mPFC and superior parietal cortex and the left ACC/mPFC and right lateral temporal cortex, another DMN subnetwork region. These results indicate the presence of CBT specific neural substrates of treatment prediction, and again, that lower baseline information transfer between these regions is indicative of greater treatment efficacy. In addition, these findings are in line with a previous study of CBT for social anxiety disorder, where increased baseline activation of the ACC and lateral temporal cortex (Klumpp et al., 2013a) was predictive of treatment response.

Central to the diffusion impairment of SN1 is a subregion of the right ACC/mPFC cortical region, area 8BM, the caudal aspect of the dorsomedial PFC (dmPFC), which borders the dorsal aspect of the ACC. Retrograde tracing studied in the macaque monkey have revealed both afferent and efferent neuronal connections between 8BM and the ACC, indicating likely functional synergy of these areas (Morecraft et al., 2012; Eradath et al., 2015). Importantly, 8BM is brain region that has recently been found to be a key region of the multiple-demand subnetwork (MDN), a fronto-parietal system that is co-activated during a broad range of cognitively demanding tasks (Assem et al., 2020). As such, the MDN is likely critical for the organization and recruitment of multiple task-specific subsystems in the brain towards current cognitive needs. Additionally, our network-diffusion model identified the most effective brain regions as putative targets for neuromodulatory stimulation on a subject-specific level. Examining the results of this investigation provide evidence that the right ACC/mPFC is an optimal target for neuromodulation to correct SN1 information diffusion dynamics of PT towards those observed in HC, and, further, that area 8BM is the most efficient subregion of the ACC/mPFC for such an intervention.

These preliminary findings have important clinical implications for the therapeutic application of neuromodulatory interventions for IPs, such as repetitive transcranial magnetic stimulation (rTMS), a focal, non-invasive brain stimulation method. rTMS, which is typically delivered to the dorsolateral PFC (dlPFC), is a first line treatment for SSRI-refractory MDD (George et al., 2013) but has also been shown to significantly reduce anxiety (Chen et al., 2019; Du et al., 2018) and nicotine dependence symptoms (Abdelrahman et al., 2021). Although rTMS is generally effective, MDD remission following treatment is 30-40% (George et al., 2013), indicating a need for developing better therapeutic paradigms, such as personalized treatment protocols. In line with these goals, a study that used MRI guidance to enhance targeting of specific regions of the dlPFC, all rTMS-resistant MDD PTs responded to this new approach (Moreno-Ortega et al., 2020). Another study used fMRI guidance to individually targeted cortical emotion regulation systems to improve rTMS efficacy (Luber et al., 2017). Our results offer an additional strategy for identifying brain network targets of rTMS treatment for correcting subject-specific structural deficits that may underlie functional expression of IP symptoms.

### Limitations and Future Directions

A significant question about the implications of our findings is whether the addition and propagation of heat on a subnetwork sufficiently models the effects of rTMS stimulation. It has been shown that the propagation of direct cortical electrode (Stiso et al., 2019) and rTMS stimulation and subsequent functional activity are best predicted by structural connectome topology (Momi et al., 2021; Beynel et al., 2020), providing encouraging evidence that network diffusion-based analyses such as those proposed in this study are an appropriate model for the effects of rTMS neuromodulation. Nevertheless, further studies are necessary to verify the utility of the proposed model.

While this is the first report of using a multimodal information diffusion model towards a transdiagnostic sample of IPs, the findings presented in this paper pertaining to the pattern of diffusion impairment of SN1 should be replicated in a larger sample of IP PT and HC to determine the generalizability of our results. This is especially true for the analyses obtained by segregating PT into SSRI and CBT cohorts, as the sample sizes of these groups are very small, and, accordingly, these findings must be interpreted as preliminary and exploratory. Further, availability of post-treatment brain imaging would be required to make reliable conclusions about network diffusion-based substrates of treatment response, and, as such, should be included in future studies. Additionally, as the data used in this manuscript are from a clinical trial conducted in 2013, and, as such, the MRI acquisition parameters result in data that is not matched in quality to that found in current high temporal and spatial resolution imaging studies, such as those following Human Connectome Project-style data collection guidelines (Glasser et al., 2013; Uğurbil et al., 2013; Sotiropoulos et al., 2013; Smith et al., 2013).

Of note, the analyses in present study are performed using a volumetric version of the HCP-MMP1.0 parcellation which has been shown to be less accurate than surface-based approaches (Coalson et al., 2018), introducing uncertainty regarding the anatomic localization of brain regions across subjects. We determined the voxel accuracy of the right area 8BM in the volumetric atlas to be 81.5% consistent with individual subject surface-derived parcellations and have otherwise limited out investigation of individual brain regions to a much coarser parcellation by aggregating HCP-MMP1.0 ROIs by their cortical label. Nonetheless, the reliability of ROI localization would be improved by using the methodology as described in (Glasser et al., 2013) and future work in which cortical-surface derived parcellations are used should adhere to these procedures.

Although our structural to functional mapping quality, as measured by Pearson correlation between the predicted and empirical functional connectomes, is in the range of values obtained in a replication study of related mappings, other network-diffusion related models have been developed that result in better predictive mappings (Deslauriers-Gauthier et al., 2020). Future studies of these proposed methods should investigate how the mapping parameters from these related methods can be incorporated into the currently proposed model for better structural-functional fusion. Another area for further development in our proposed network diffusion methods is that neuromodulation target identification and simulation can only model the effects of the *addition* of heat to subnetwork’s diffusion dynamics. Therefore, if a brain circuit is found to be pathologically hyperconnected, other strategies will have to be developed in order model either direct or indirect inhibition of brain activity. Furthermore, while the network-diffusion approaches used in this study have been used to successfully demonstrate the intimate relationship between structural and functional brain connectivity, they do not model directed communication between brain regions. Future analyses could potentially incorporate findings from anatomical tracing studies such that reconstructed fibers corresponding to such well-characterized white matter tracts between ROIs may be assigned a direction of signal propagation. Regardless, advances in network analyses that consider the inherently directed nature of neural communication are needed to in order to more accurately model the underlying neurobiology of brain structure and function.

Future implications of this research are broad. We present here an analytical framework assembled from well-characterized neuroimaging and graph theoretical methods that can be used as is to study multimodal brain networks for other brain disorders. Aside from the application to other datasets and to larger similar datasets for replication, this model offers the ability to determine the diffusion embedding from functional connectivity other than that observed during resting-state. For example, functional connectomes can be generated from tasks that are known to be implicated in a certain disorder, and as a result the diffusion embedding will be formed from the structural basis that best maps to the functional connectivity observed during this task. This allows for the researcher to investigate the structural connectivity potentially most pertinent to forming the functional brain activity observed during specific tasks, and therefore permits a more granular interrogation of complex features of brain disorders and states. Perhaps the most promising and immediate application of our proposed methodology is that of identifying brain regions best suited for therapeutic for neuromodulatory intervention, such as with rTMS. As the proposed methods provide both subject-specific regional targets and magnitudes of stimulation that best modify subnetwork dynamics towards those of a desired (HC) network, future work is in line with improving the outcomes of rTMS intervention by personalizing treatment features.

## Conclusion

There are two major outcomes of this study. First, we report impaired information diffusion between area 8BM and other SN1 regions, many of which have been previously implicated in both the pathology of multiple IPs and the function of the default mode, cingulo-opercular and dorsal affective networks. Second, we found that hubs of SN1, found to be critical for organizing brain function for a variety of cognitive and emotional processes, are optimal targets for modelled neuromodulatory intervention. Taken together, our results may indicate the presence of a concerted disruption of multiple brain networks pertinent to cognitive control of emotion regulation in IPs. This dysregulation of connectivity could result in a loss of “top-down” executive control of emotion processing via connections between the multiple demand network and other task-specific (and task-negative) brain networks. Such a perturbation in brain network dynamics involving the integration of multiple complex subsystems via a hub of the multiple demand network may represent the underlying pathophysiological brain network features that are common to all IPs and give rise to the heterogeneous expression of transdiagnostic symptoms.

## Data Availability

Data and software specific to this manuscript are available at the lead author's github page. All other resources are detailed in the methods section of the manuscript.

https://github.com/pauljasonthomas/brainnetdiff

## ACKNOWLEDGEMENTS

This work was supported by funding from the National Institute of Mental Health of the National Institutes of Health (NIMH-NIH) grants R01MH101497 (to KLP) and 5T32MH067631-14 (support for PJT).

## Notes

Data and code used in this manuscript can be found at the lead author’s github: https://github.com/pauljasonthomas/brainnetdiff.

