## Supplemental Information for "Network Diffusion Embedding Reveals Transdiagnostic Subnetwork Disruption and Potential Treatment Targets in Internalizing Psychopathologies"

#### GRAPH THEORETICAL ANALYSIS OF STRUCTURAL CONNECTOMES

In order to determine the relationship between the structural diffusion distance (SDD) connectomes proposed in this study and canonical measures of network topology, we computed the nodal strength, betweenness centrality, local efficiency and clustering coefficent for each ROI individually and meaned by cortical region both using the entire brain network (global) and the subgraph of nodes (local) found only in SN1 (Figure S1). We found that the hubbness of regions in SDD connectomes generally did not align with that found in the global or local SN1 regions, especially pertaining to the right area 8BM, indicating that the SDD connectomes capture information that is not manifestly present in typical adjacency matrices of structural connectivity.

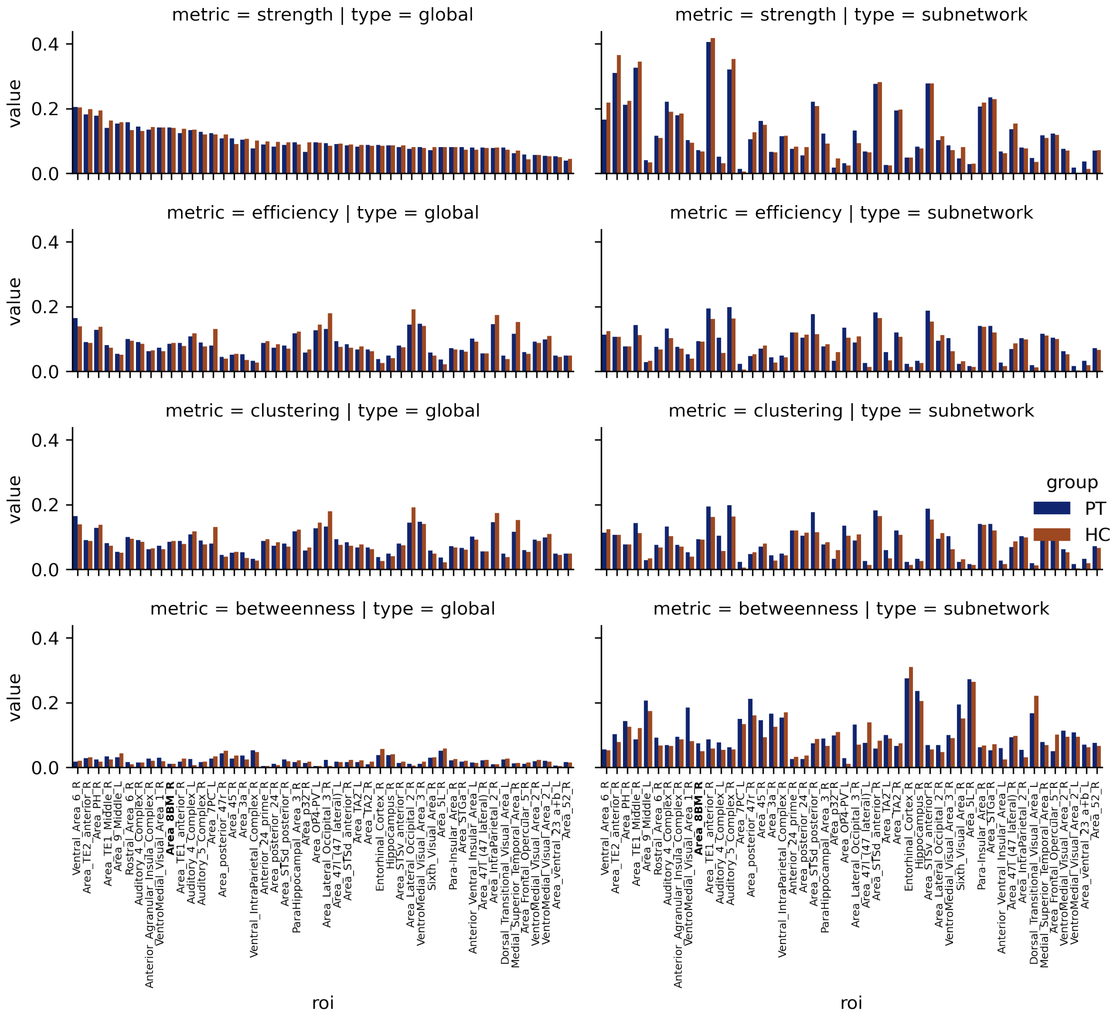


**Figure S1:** Average graph theory metrics computed for HC and PT for nodes of SN1 in the whole brain subnetwork (global, first column) or in the subgraph of SN1 only (subnetwork, second column). Metrics computed are strength, local efficiency, clustering coefficient and betweenness centrality from top to bottom row, respectively. Note that the label for the bars corresponding to the right area 8BM are bolded.

#### ANALYSIS OF VOLUMETRIC HCP-MMP1.0 PARCELLATION

A surface to volume projected version of the HCP-MMP1.0 atlas (Glasser et al., 2016) registered to MNI space and corrected for errors arising from surface-voxel misalignment (as implemented in DSI Studio) was used for the present analyses. To determine the localization of the right area 8BM parcel from the volumetric HCP-MMP1.0, we determined the percentage of voxels that are labeled by ROIs from derived from individual subject surface-registered HCP-MMP1.0 parcellations mapped to MNI volumetric space.

##### Surface to volume mapping

First, cortical reconstruction of the T1w structural imaging data was performed with the FreeSurfer Image analysis suite (http://surfer.nmr.mgh.harvard.edu/) using the automated procedures in the recon-all pipeline, described in (Dale et al., 1999; Fischl et al., 2002). In brief, the structural images underwent the removal of non-brain tissue (Ségonne et al., 2004), transformation to Talairach space, segmentation of white and gray matter structures (Fischl et al., 2002, 2004), intensity normalization (Sled et al., 1998), tessellation of gray-white matter boundary, topological correction (Fischl et al., 2001; Ségonne et al., 2007) and surface deformation using intensity gradients for optimal placement of the white-gray matter boundary (Dale et al., 1999; Dale and Sereno, 1993; Fischl and Dale, 2000). Once complete, cortical surface models were inflated (Fischl et al., 1999a) and registered to a spherical atlas (Fischl et al., 1999b). Next, following the "Resampling FreeSurfer" guide (available at https://wiki.humanconnectome.org/display/PublicData/HCP+Users+FAQ#HCPUsersFAQ-9.HowdoImapdatabetweenFreeSurferandHCP? surface data (FreeSurfer pial, white and sphere.reg files) were resampled to the 32k fs_LR mesh on which the HCP-MMP1.0 parcellation resides using the Connectome Workbench (Glasser et al., 2013) workbench commands wb_command -freesurfer-resample-prep and wb_command -surface-resample. We then mapped the HCP-MMP1.0 parcellation from each subject's surface mesh to their native volume space using a ribbon constrained mapping algorithm as implemented in the workbench command wb_command -label-to-volume-mapping. Lastly, T1w volumes were transformed to MNI space using the nonlinear registration method implemented in DSI Studio (using the command dsi_studio –action=reg) and the resulting warpfield was used to transform MMP-HCP1.0 native volume parcellations to MNI space.

**Localization of right area 8BM**

Once we had obtained subject-specific surface registered HCP-MMP1.0 parcellations mapped to standardized MNI volumetric space, we used these data to investigate the anatomic localization of the right area 8BM. We first created a voxel probability map to determine the of anatomic localization of the surface to volume mapped right area 8BM parcel compared to that of the MNI template volumetric atlas used to define ROIs in the present study (Figure S2A). In this visualization, each voxel value corresponds to the number of subjects in which the surface to volume mapped parcellation labels it as the right area 8BM divided by the total number of subjects. Although this procedure demonstrated satisfactory visual correspondence between the two parcellation approaches, we next sought quantify the consistency of parcel localization. To do this, we calculated the quantity of individual subject surface-registered right 8BM parcel voxels that are mapped to each MNI template registered ROI. In doing this we found that 81.5% of surface to volume derived right 8BM parcel voxels correctly corresponded to the MNI registered template volume (Figure S2B). The majority of incorrectly labeled MNI template registered parcellation voxels were found in the neighboring right SFL, SCEF and 8BL parcels (6.6%, 5.4% and 5.0%, respectively).

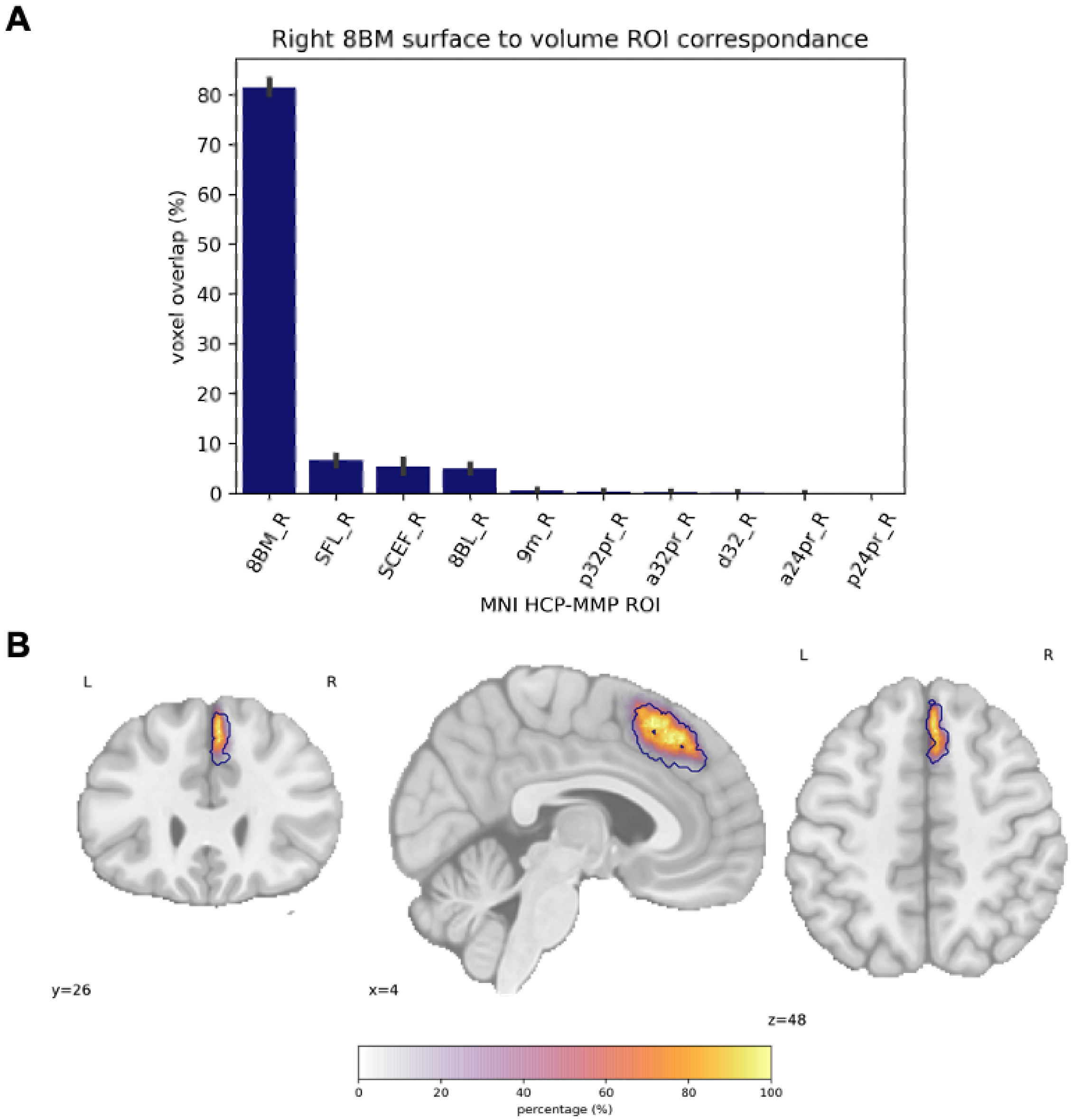


**Figure S2:** Consistency of anatomic localization of right area 8BM in the MNI registered volumetric HCP-MMP1.0 atlas. **A** Probability map of surface to volume mapped right 8BM parcel voxels compared to MNI template registered right 8BM (dark blue outline). Voxel values are determined by the number of subjects in which a voxel is labeled as right 8BM via their surface to volume mapped parcel, divided by the total number of subject. **B** Bar plot visualization of the percentage of surface to volume right 8BM parcel voxels correspondence to MNI registered template ROIs. Note that only ROIs with voxel percentage $> 0\%$ are shown.

### HEAT KERNEL VALUE CORRELATIONS WITH IDAS-II SCALES

Baseline Depression (all PT)

| roi1 | roi2 | rho | p value | q value |
|---|---|---|---|---|
| Anterior_Cingulate_and_Medial_Prefrontal_L | Posterior_Cingulate_L | 0.393 | 0.001 | 0.036 |
| Anterior_Cingulate_and_Medial_Prefrontal_R | Inferior_Frontal_L | -0.365 | 0.003 | 0.036 |
| Insular_and_Frontal_Opercular_L | Anterior_Cingulate_and_Medial_Prefrontal_L | -0.356 | 0.004 | 0.036 |
| Inferior_Frontal_L | Anterior_Cingulate_and_Medial_Prefrontal_L | -0.354 | 0.004 | 0.036 |
| Dorsal_Stream_Visual_R | Anterior_Cingulate_and_Medial_Prefrontal_L | 0.351 | 0.004 | 0.036 |
| Ventral_Stream_Visual_L | Anterior_Cingulate_and_Medial_Prefrontal_L | 0.340 | 0.006 | 0.040 |
| Anterior_Cingulate_and_Medial_Prefrontal_R | Insular_and_Frontal_Opercular_L | -0.318 | 0.010 | 0.061 |
| Inferior_Frontal_R | Anterior_Cingulate_and_Medial_Prefrontal_L | -0.283 | 0.022 | 0.115 |
| Insular_and_Frontal_Opercular_R | Anterior_Cingulate_and_Medial_Prefrontal_R | -0.280 | 0.024 | 0.115 |
| Inferior_Frontal_R | Anterior_Cingulate_and_Medial_Prefrontal_R | -0.269 | 0.030 | 0.131 |
| Medial_Temporal_R | Anterior_Cingulate_and_Medial_Prefrontal_L | 0.245 | 0.049 | 0.191 |

Depression treatment response (all PT)

| roi1 | roi2 | rho | p value | q value |
|---|---|---|---|---|
| Inferior_Frontal_L | Anterior_Cingulate_and_Medial_Prefrontal_L | -0.450 | 0.001 | 0.042 |
| Auditory_Association_L | Anterior_Cingulate_and_Medial_Prefrontal_L | -0.421 | 0.002 | 0.042 |
| Insular_and_Frontal_Opercular_L | Anterior_Cingulate_and_Medial_Prefrontal_L | -0.412 | 0.003 | 0.042 |
| Anterior_Cingulate_and_Medial_Prefrontal_R | Inferior_Frontal_L | -0.388 | 0.005 | 0.057 |
| Inferior_Frontal_R | Anterior_Cingulate_and_Medial_Prefrontal_L | -0.373 | 0.008 | 0.059 |
| Anterior_Cingulate_and_Medial_Prefrontal_R | Auditory_Association_L | -0.369 | 0.008 | 0.059 |
| Lateral_Temporal_R | Anterior_Cingulate_and_Medial_Prefrontal_L | -0.334 | 0.018 | 0.099 |
| Lateral_Temporal_R | Anterior_Cingulate_and_Medial_Prefrontal_R | -0.332 | 0.018 | 0.099 |
| Inferior_Frontal_R | Anterior_Cingulate_and_Medial_Prefrontal_R | -0.325 | 0.021 | 0.101 |
| Insular_and_Frontal_Opercular_R | Anterior_Cingulate_and_Medial_Prefrontal_L | -0.308 | 0.030 | 0.118 |
| Insular_and_Frontal_Opercular_R | Anterior_Cingulate_and_Medial_Prefrontal_R | -0.305 | 0.031 | 0.118 |
| Anterior_Cingulate_and_Medial_Prefrontal_R | Superior_Parietal_R | -0.302 | 0.033 | 0.118 |
| Anterior_Cingulate_and_Medial_Prefrontal_R | Insular_and_Frontal_Opercular_L | -0.290 | 0.041 | 0.135 |

Panic treatment response (SSRI only)

| roi1 | roi2 | rho | p value | q value |
|---|---|---|---|---|
| Inferior_Frontal_L | Anterior_Cingulate_and_Medial_Prefrontal_L | -0.596 | 0.003 | 0.147 |
| Insular_and_Frontal_Opercular_L | Anterior_Cingulate_and_Medial_Prefrontal_L | -0.543 | 0.009 | 0.174 |
| Insular_and_Frontal_Opercular_R | Anterior_Cingulate_and_Medial_Prefrontal_L | -0.525 | 0.012 | 0.174 |
| Inferior_Frontal_R | Anterior_Cingulate_and_Medial_Prefrontal_L | -0.498 | 0.018 | 0.197 |
| Lateral_Temporal_R | Anterior_Cingulate_and_Medial_Prefrontal_L | -0.465 | 0.029 | 0.250 |

Depression treatment response (CBT only)

| roi1 | roi2 | rho | p value | q value |
|---|---|---|---|---|
| Anterior_Cingulate_and_Medial_Prefrontal_R | Superior_Parietal_R | -0.507 | 0.006 | 0.082 |
| Inferior_Parietal_R | Anterior_Cingulate_and_Medial_Prefrontal_R | -0.506 | 0.006 | 0.082 |
| Anterior_Cingulate_and_Medial_Prefrontal_R | Auditory_Association_L | -0.492 | 0.008 | 0.082 |
| Anterior_Cingulate_and_Medial_Prefrontal_R | Inferior_Frontal_L | -0.489 | 0.008 | 0.082 |
| Anterior_Cingulate_and_Medial_Prefrontal_R | Insular_and_Frontal_Opercular_L | -0.481 | 0.009 | 0.082 |
| Auditory_Association_L | Anterior_Cingulate_and_Medial_Prefrontal_L | -0.463 | 0.013 | 0.084 |
| Inferior_Parietal_R | Anterior_Cingulate_and_Medial_Prefrontal_L | -0.449 | 0.017 | 0.084 |
| Lateral_Temporal_R | Anterior_Cingulate_and_Medial_Prefrontal_R | -0.449 | 0.017 | 0.084 |
| Insular_and_Frontal_Opercular_L | Anterior_Cingulate_and_Medial_Prefrontal_L | -0.446 | 0.017 | 0.084 |
| Anterior_Cingulate_and_Medial_Prefrontal_R | MT+_Complex_and_Neighboring_Visual_Areas_R | -0.399 | 0.035 | 0.152 |

Panic treatment response (CBT only)

| roi1 | roi2 | rho | p value | q value |
|---|---|---|---|---|
| Anterior_Cingulate_and_Medial_Prefrontal_R | Superior_Parietal_R | -0.545 | 0.003 | 0.117 |
| Lateral_Temporal_R | Anterior_Cingulate_and_Medial_Prefrontal_L | -0.500 | 0.007 | 0.144 |
| Inferior_Frontal_L | Anterior_Cingulate_and_Medial_Prefrontal_L | -0.414 | 0.028 | 0.370 |
| Insular_and_Frontal_Opercular_L | Anterior_Cingulate_and_Medial_Prefrontal_L | -0.401 | 0.034 | 0.370 |

**Table S1** Correlations between either baseline or percentage reduction following treatment (treatment response) of IDAS-II Panic and Depression subscales and heat kernel values. Columns 'roi1' and 'roi2' indicate SN1 brain regions, while columns 'rho', 'p value' and 'q value' indicate the Spearman rho statistic, corresponding uncorrected p-value and and FDR corrected p-value, respectively. Data in this table corresponds to the analyses illustrated in Figure 3 of the main text. All significant correlations before FDR correction are shown.

#### BASELINE AND POST-HEAT SUPPLEMENT HEAT KERNEL VALUES

| | | baseline | | | post heat supplement | | |
|---|---|---|---|---|---|---|---|
| roi1 | roi2 | t statistic | p value | q value | t statistic | p value | q value |
| Paracentral_Lobular_and_Mid_Cingulate_R | Inferior_Frontal_L | 3.493 | 0.001 | 0.023 | 2.933 | 0.004 | 0.086 |
| Paracentral_Lobular_and_Mid_Cingulate_R | Insular_and_Frontal_Opercular_L | 3.380 | 0.001 | 0.023 | 2.548 | 0.013 | 0.154 |
| Somatosensory_and_Motor_R | Inferior_Frontal_L | 3.068 | 0.003 | 0.036 | 2.221 | 0.029 | 0.210 |
| Somatosensory_and_Motor_R | Dorsal_Stream_Visual_R | 3.261 | 0.002 | 0.030 | 2.721 | 0.008 | 0.125 |
| Premotor_R | Dorsal_Stream_Visual_R | 3.377 | 0.001 | 0.023 | 3.107 | 0.003 | 0.072 |
| Anterior_Cingulate_and_Medial_Prefrontal_R | Posterior_Cingulate_L | 3.382 | 0.001 | 0.023 | 2.238 | 0.028 | 0.210 |
| Anterior_Cingulate_and_Medial_Prefrontal_R | Ventral_Stream_Visual_L | 1.968 | 0.052 | 0.229 | -3.836 | 0.000 | 0.027 |
| Early_Auditory_R | Anterior_Cingulate_and_Medial_Prefrontal_L | 3.530 | 0.001 | 0.023 | 0.362 | 0.718 | 0.871 |
| Early_Auditory_R | Inferior_Frontal_L | 3.042 | 0.003 | 0.036 | 2.182 | 0.032 | 0.218 |
| Early_Auditory_R | Insular_and_Frontal_Opercular_L | 3.247 | 0.002 | 0.030 | 2.264 | 0.026 | 0.210 |
| Early_Auditory_R | Dorsal_Stream_Visual_R | 2.956 | 0.004 | 0.043 | 2.893 | 0.005 | 0.087 |
| Early_Auditory_R | Anterior_Cingulate_and_Medial_Prefrontal_R | 3.736 | 0.000 | 0.023 | -3.091 | 0.003 | 0.072 |
| Auditory_Association_R | Anterior_Cingulate_and_Medial_Prefrontal_L | 3.754 | 0.000 | 0.023 | 3.213 | 0.002 | 0.072 |
| Auditory_Association_R | Dorsal_Stream_Visual_R | 3.506 | 0.001 | 0.023 | 3.483 | 0.001 | 0.040 |
| Auditory_Association_R | Anterior_Cingulate_and_Medial_Prefrontal_R | 3.565 | 0.001 | 0.023 | 2.374 | 0.020 | 0.186 |
| Insular_and_Frontal_Opercular_R | Posterior_Cingulate_L | 3.188 | 0.002 | 0.034 | 2.962 | 0.004 | 0.086 |
| Insular_and_Frontal_Opercular_R | Dorsal_Stream_Visual_R | 3.803 | 0.000 | 0.023 | 3.748 | 0.000 | 0.027 |
| Lateral_Temporal_R | Anterior_Cingulate_and_Medial_Prefrontal_L | 3.401 | 0.001 | 0.023 | 2.484 | 0.015 | 0.160 |
| Lateral_Temporal_R | Dorsal_Stream_Visual_R | 3.636 | 0.000 | 0.023 | 3.608 | 0.001 | 0.033 |
| Lateral_Temporal_R | Anterior_Cingulate_and_Medial_Prefrontal_R | 3.029 | 0.003 | 0.036 | 0.836 | 0.406 | 0.772 |
| Inferior_Parietal_R | MT+_Complex_and_Neighboring_Visual_Areas_R | 3.125 | 0.002 | 0.036 | 3.108 | 0.003 | 0.072 |
| Inferior_Parietal_R | Anterior_Cingulate_and_Medial_Prefrontal_R | 1.114 | 0.268 | 0.595 | -4.098 | 0.000 | 0.024 |
| Inferior_Parietal_R | Lateral_Temporal_R | 3.089 | 0.003 | 0.036 | 3.075 | 0.003 | 0.072 |
| Ventral_Stream_Visual_R | Anterior_Cingulate_and_Medial_Prefrontal_L | 3.066 | 0.003 | 0.036 | 1.856 | 0.067 | 0.326 |
| Ventral_Stream_Visual_R | Anterior_Cingulate_and_Medial_Prefrontal_R | 3.169 | 0.002 | 0.034 | 0.661 | 0.511 | 0.828 |
| Ventral_Stream_Visual_R | Inferior_Parietal_R | 3.042 | 0.003 | 0.036 | 2.840 | 0.006 | 0.095 |

**Table S2:** T-test results for mean heat kernel values (HKVs) at baseline and after supplemental heat (post heat supplement) in healthy controls versus patients. HKVs of brain region pairs, indicated by columns 'roi1' and 'roi2', are mean cortical regions of SN1 defined by the MMP1.0 parcellation. Data in this table corresponds to the analyses illustrated in Figure 4 of the main text. The 'q value' column contains FDR corrected p-values.

| | | baseline | | | post heat supplement | | |
|---|---|---|---|---|---|---|---|
| roi1 | roi2 | t statistic | p value | q value | t statistic | p value | q value |
| Area__5L__R | Area__47l__(47__lateral)__L | 3.493 | 0.001 | 0.045 | 3.240 | 0.002 | 0.089 |
| Area__5L__R | Anterior__Ventral__Insular__Area__L | 3.380 | 0.001 | 0.046 | 2.991 | 0.004 | 0.097 |
| Ventral__Area__6__R | Sixth__Visual__Area__R | 3.571 | 0.001 | 0.045 | 3.445 | 0.001 | 0.063 |
| Area__8BM__R | VentroMedial__Visual__Area__2__L | 1.491 | 0.140 | 0.392 | -4.211 | 0.000 | 0.014 |
| Area__8BM__R | Area__Lateral__Occipital__2__R | 2.973 | 0.004 | 0.059 | -6.472 | 0.000 | 0.000 |
| Area__p32__R | Area__ventral__23__a+b__L | 4.144 | 0.000 | 0.032 | 4.010 | 0.000 | 0.024 |
| Area__p32__R | Dorsal__Transitional__Visual__Area__L | 3.409 | 0.001 | 0.046 | 2.914 | 0.005 | 0.109 |
| Area__52__R | Area__9__Middle__L | 3.530 | 0.001 | 0.045 | 2.090 | 0.040 | 0.308 |
| Area__52__R | Anterior__24__prime__R | 3.521 | 0.001 | 0.045 | 0.740 | 0.461 | 0.788 |
| Area__52__R | Area__8BM__R | 3.361 | 0.001 | 0.046 | -3.203 | 0.002 | 0.094 |
| Area__52__R | Area__p32__R | 3.545 | 0.001 | 0.045 | 2.759 | 0.007 | 0.129 |
| Area__TA2__R | Area__9__Middle__L | 4.080 | 0.000 | 0.032 | 2.789 | 0.007 | 0.125 |
| Area__TA2__R | Area__8BM__R | 3.441 | 0.001 | 0.046 | -1.904 | 0.060 | 0.344 |
| Area__TA2__R | Area__p32__R | 3.476 | 0.001 | 0.045 | 2.827 | 0.006 | 0.125 |
| Anterior__Agranular__Insula__Complex__R | Dorsal__Transitional__Visual__Area__L | 3.857 | 0.000 | 0.036 | 3.789 | 0.000 | 0.032 |
| Anterior__Agranular__Insula__Complex__R | Sixth__Visual__Area__R | 3.938 | 0.000 | 0.032 | 3.900 | 0.000 | 0.024 |
| Auditory__5__Complex__R | Area__9__Middle__L | 4.108 | 0.000 | 0.032 | 2.897 | 0.005 | 0.112 |
| Auditory__5__Complex__R | Area__p32__R | 3.608 | 0.001 | 0.045 | 2.978 | 0.004 | 0.097 |
| Area__STSd__anterior__R | Sixth__Visual__Area__R | 3.590 | 0.001 | 0.045 | 3.578 | 0.001 | 0.054 |
| Area__STSd__anterior__R | Area__8BM__R | 3.408 | 0.001 | 0.046 | -2.374 | 0.020 | 0.219 |
| Area__STSd__posterior__R | Sixth__Visual__Area__R | 3.730 | 0.000 | 0.045 | 3.709 | 0.000 | 0.038 |
| Area__TE1__anterior__R | Sixth__Visual__Area__R | 3.946 | 0.000 | 0.032 | 3.931 | 0.000 | 0.024 |
| Area__TE2__anterior__R | Area__9__Middle__L | 4.046 | 0.000 | 0.032 | 3.066 | 0.003 | 0.097 |
| Area__TE2__anterior__R | Sixth__Visual__Area__R | 3.371 | 0.001 | 0.046 | 3.359 | 0.001 | 0.070 |
| Area__TE2__anterior__R | Area__8BM__R | 3.585 | 0.001 | 0.045 | -1.595 | 0.114 | 0.451 |
| Area__IntraParietal__2__R | Area__8BM__R | 0.269 | 0.789 | 0.899 | -3.921 | 0.000 | 0.024 |
| VentroMedial__Visual__Area__1__R | Area__p32__R | 3.533 | 0.001 | 0.045 | 3.089 | 0.003 | 0.097 |
| VentroMedial__Visual__Area__3__R | Area__8BM__R | 2.863 | 0.005 | 0.069 | -4.648 | 0.000 | 0.004 |
| Area__Lateral__Occipital__3__R | Area__8BM__R | 1.614 | 0.110 | 0.357 | -4.588 | 0.000 | 0.004 |
| VentroMedial__Visual__Area__2__R | Area__8BM__R | 3.104 | 0.003 | 0.057 | -4.823 | 0.000 | 0.003 |
| Auditory__4__Complex__R | Area__9__Middle__L | 3.336 | 0.001 | 0.046 | 2.241 | 0.028 | 0.261 |
| Auditory__4__Complex__R | Area__47l__(47__lateral)__L | 3.493 | 0.001 | 0.045 | 3.184 | 0.002 | 0.096 |
| Auditory__4__Complex__R | Anterior__Ventral__Insular__Area__L | 3.339 | 0.001 | 0.046 | 2.990 | 0.004 | 0.097 |
| Auditory__4__Complex__R | Area__p32__R | 3.715 | 0.000 | 0.045 | 3.067 | 0.003 | 0.097 |
| Area__STSv__anterior__R | Sixth__Visual__Area__R | 3.421 | 0.001 | 0.046 | 3.401 | 0.001 | 0.067 |
| Para-Insular__Area__R | Sixth__Visual__Area__R | 3.348 | 0.001 | 0.046 | 3.336 | 0.001 | 0.071 |
| Area__posterior__24__R | Area__52__R | 3.370 | 0.001 | 0.046 | 1.337 | 0.185 | 0.534 |

**Table S3:** T-test results for mean heat kernel values (HKVs) at baseline and after supplemental heat (post heat supplement) in healthy controls versus patients. HKVs of brain region pairs, indicated by columns 'roi1' and 'roi2', are individual regions of interest (ROIs) of SN1 defined by the MMP1.0 parcellation. Data in this table corresponds to the analyses illustrated in Figure 4 of the main text. The 'q value' column contains FDR corrected p-values.
